# Efficacy and Safety of a Plant-Based Virus-Like Particle Vaccine for COVID-19 Adjuvanted with AS03

**DOI:** 10.1101/2022.01.17.22269242

**Authors:** Karen Joyce Hager, Gonzalo Pérez Marc, Philipe Gobeil, Ricardo Sobhie Diaz, Gretchen Heizer, Conrado Llapur, Alexander I. Makarkov, Eduardo Vasconcellos, Stephane Pillet, Fernando Riera, Kapil Bhutada, Priscila Geller Wolff, Garry Wallace, Hessam Aazami, Christine E. Jones, Fernando P. Polack, Judith Atkins, Iohann Boulay, Jiwanjeet Dhaliwall, Nathalie Charland, Manon Couture, Julia Jiang-Wright, Nathalie Landry, Sophie Lapointe, Aurélien Lorin, Asif Mahmood, Lawrence H. Moulton, Emmy Pahmer, Julie Parent, Pooja Saxena, Annie Séguin, Luan Tran, Thomas Breuer, Maria Angeles Ceregido, Marguerite Koutsoukos, François Roman, Junya Namba, Marc-André D’Aoust, Sonia Trepanier, Yosuke Kimura, The CoVLP Study Team, Brian J. Ward

## Abstract

**Background:** Several COVID-19 vaccines are currently being deployed but supply constraints, concerns over durability of immune responses, solidifying vaccine hesitancy/resistance and vaccine efficacy in the face of emerging variants mean that new vaccines continue to be needed to fight the ongoing pandemic. The vaccine described here is an enveloped, coronavirus-like particle produced in plants (CoVLP) that displays the prefusion-stabilized spike (S) glycoprotein of SARS-CoV-2 (ancestral Wuhan strain) and is adjuvanted with AS03 (CoVLP+AS03).

**Methods:** This Phase 3 randomized, observer-blind, placebo-controlled trial was conducted at 85 centers in Argentina, Brazil, Canada, Mexico, the UK, and the USA. Adults ≥18 years of age including those at high risk for COVID-19 complications were randomly assigned 1:1 to receive two intramuscular injections of CoVLP (3.75 μg) adjuvanted with AS03 or placebo, 21 days apart. The primary efficacy endpoint was prevention of symptomatic (≥ 1 symptom), PCR-confirmed SARS-CoV-2 infection with onset at least 7 days after the second injection and was triggered by the identification of ≥160 virologically-confirmed cases. Tolerability and safety of CoVLP+AS03 were also determined.

**Results:** A total of 24,141 volunteers were randomly assigned 1:1 to receive vaccine or placebo (N= 12,074 and 12,067, respectively: median age 29, range 18 to 86 years). Overall, 83% received both doses. 14.8% were SARS-CoV-2 seropositive at baseline. Symptomatic SARS-CoV-2 infection was confirmed in 165 study participants in the intention to treat (ITT) set and 157 in the per-protocol population (PP) set. Of the 157 in the PP set, 118 COVID-19 cases were in the placebo group and 39 COVID-19 cases were in the CoVLP+AS03 group for an overall vaccine efficacy (VE) of 71.0% (95% confidence interval (CI) 58.6, 80.0). Moderate-to-severe COVID-19 occurred in 8 and 32 participants in the CoVLP+AS03 and placebo groups, respectively: VE 78.1% (95% CI: 53.9, 90.5) in the PP set overall and 84.5% (95% CI: 62.0, 94.7) in those seronegative at recruitment.

To date, 100% of the sequenced strains (122/165 cases: 73.39%) were variants, dominated by Delta (45.9%) and Gamma (43.4%) strains. Vaccine efficacy by variant was 75.3% (95% CI 52.8, 87.9) against Delta and 88.6% (95% CI 74.6, 95.6) against Gamma. Cross-protection was also observed against Alpha, Lambda and Mu variants; although fewer cases were identified, all were in the placebo group. At diagnosis, viral loads in the CoVLP+AS03 breakthrough cases were >100-fold lower than in the placebo cases. Reactogenicity data for solicited adverse events (AEs) was analysed for a subset (N=4,136 in vaccine arm and N=3,683 for placebo) of participants. Reactogenicity was mostly mild to moderate, and transient, and occurred more frequently in the CoVLP+AS03 group. The safety analysis set used for unsolicited AE assessment comprised 24,076 participants who received at least one study injection: 12,036 received CoVLP+AS03 and 12,040 received placebo. All serious adverse events were assessed as unrelated, except two events reported in the same subject in the placebo group. No significant imbalance or safety concern was noted in medically attended AEs (MAAEs), adverse event of special interest (AESIs), AEs leading to withdrawal, deaths, or adverse events potentially associated with currently authorized vaccines.

**Conclusions:** The CoVLP+AS03 vaccine candidate conferred an efficacy of 71.0% in preventing symptomatic SARS-CoV-2 infection caused by a spectrum of variants. Vaccine efficacy of 78.1% was observed against moderate and severe disease, while variant-specific efficacy ranged from 75.3% to 100%. Markedly lower viral loads in the CoVLP+AS03 group at the time of diagnosis suggests a significant virologic impact of vaccination even in the breakthrough cases. CoVLP+AS03 vaccine candidate was well tolerated, and no safety concerns were identified during the study. If approved by regulators, this more traditional protein+adjuvant vaccine produced using the novel plant-based platform may be able to make an important contribution to the global struggle against the increasingly complex family of SARS-CoV-2 viruses (Funded by Medicago with grants from the governments of Quebec and Canada; NCT04636697).

## Introduction

Since its emergence in late 2019 ^1^, the severe acute respiratory syndrome coronavirus 2 (SARS-CoV-2) has caused more than 312 million cases of coronavirus disease 2019 (COVID-19) globally with >5.5 million deaths ^2^. A massive global effort began almost immediately upon recognition of the new virus that resulted in the development of a large diversity of vaccine candidates based on mRNA, non-replicating adenovirus-vectored, attenuated and inactivated viruses, and adjuvanted protein-based platforms ^3^. The spike (S) glycoprotein contains a receptor binding domain (RBD) that binds to human angiotensin-converting enzyme 2 (ACE2) receptor which initiates viral fusion and entry into host cells ^4,5^. Neutralizing antibodies (NAb) directed against the S protein provide protection from other highly pathogenic coronaviruses (e.g.: SARS-CoV-1, MERS) and similar protective efficacy was rapidly demonstrated with anti-S antibodies in SARS-CoV-2 infection ^6^. Consequently, the S protein is the target for almost all COVID-19 vaccines except for attenuated and inactivated virus-based vaccine candidates. Phase 3 clinical trials launched during the first waves of the pandemic generally demonstrated high vaccine efficacy against the ancestral (Wuhan) SARS-CoV-2 strain ^7^. Several vaccines have since been deployed around the world with considerable success ^8-10^ and acceptable safety profiles despite some concerns regarding rare cases of myocarditis and pericarditis after mRNA vaccines ^11^ and thrombotic events associated with some adenovirus vectored vaccines ^12^ which have been added to the prescribing information. However, the reduced protection reported more recently, from clinical trials and subsequent real-world data ^13-17^, may be due to a combination of reduced cross-reactivity of vaccine-induced antibodies against emerging SARS-CoV-2 variants and waning vaccine-induced humoral immunity with time, especially for mild disease ^18,19^. A third dose (booster) of the mRNA vaccines has been reported to restore serum NAb levels and improve cross-protection against emerging variants, especially against the most severe outcomes ^20^. Tensions created by the simultaneous demand for booster doses in countries with largely immunized populations and the need to provide primary vaccination to the majority of the world’s unvaccinated populations ^6,21,22^ emphasize the need for additional vaccines and vaccine suppliers to fully meet the global demand. Alternative vaccine options that can be used under routine storage and handling conditions (2-8°C) or that can overcome the concerns of individuals who hesitate to get vaccinated with currently licensed vaccines based on novel platform technologies due to medical conditions or beliefs ^23^ would also be useful.

Virus-like particles (VLP)-based vaccine have been highly successful against several viral pathogens such as the Hepatitis B virus and the Human Papillomavirus ^24^. Medicago has developed a plant-based production platform to generate VLPs for a range of viral pathogens including enveloped VLP candidates for pandemic and seasonal influenza that have demonstrated substantial immunogenicity or efficacy in human studies ^25-29^. This vaccine manufacturing platform uses transient transfection of *Nicotiana benthamiana*, a common Australian plant and a disarmed *Agrobacterium tumefaciens* vector to deliver the episomal DNA encoding the vaccine protein to the plant cell nucleus ^30^. Expression of the full-length, pre-fusion-stabilized SARS-CoV-2 S protein (ancestral Wuhan strain) in plant cells results in the spontaneous formation of 100-150 nm enveloped VLPs (hereafter referred to as CoVLP). Once purified, CoVLP is stable for at least 6 months under routine vaccine storage/transportation conditions (2-8°C). The Adjuvant System 03 (AS03) is an established adjuvant manufactured by GlaxoSmithKline (GSK). AS03 initiates a transient innate response at the injection site and the draining lymph node in animal models ^31^, and in human peripheral blood cells^32-34^. This innate immune activity potentiates and shapes both the humoral and cell-mediated adaptive responses to the vaccine antigen, resulting in increased magnitude, quality (e.g.: antibody avidity), breadth and durability of the immune responses ^35-39^. AS03 has been used to adjuvant pandemic A/H1N1pdm09 influenza vaccines (>90 million doses pandemic influenza vaccines have been administered worldwide), as well as in other licensed vaccines or vaccine candidates ^40^. AS03-adjuvanted CoVLP (CoVLP+AS03) has been shown to induce strong and durable neutralizing antibody responses, as well as a balanced IFN-γ/IL-4 T cell response ^41,42^, both of which are likely to be important in protecting against SARS-CoV-2 infection ^43^.

We report herein the results of the pivotal Phase 3 portion of a Phase 2/3 study in which the safety and efficacy of CoVLP+AS03 were evaluated in 24,141 adult participants ≥18 years of age recruited between March 15^th^, 2021, and September 2^nd^, 2021, in six countries across three continents (Europe, North and South America). This placebo-controlled trial was conducted during an evolving pandemic environment with active circulation of several variants of concern and of interest, as well as increasing vaccine roll-out.

## Methods

### Trial Objectives and Oversight

The trial is being conducted in accordance with current International Conference on Harmonization (ICH) guidelines on Good Clinical Practice, and applicable country-specific regulatory requirements. All participants provided written informed consent before being enrolled. An independent Institutional Review Board (IRB) or Ethics Committee (EC) approved the protocol, protocol amendments and consent forms. Safety and efficacy data, as required by protocol, was reviewed by an Independent Data Monitoring Committee (IDMC) as needed.

The objectives of the randomized, observed-blinded, placebo-controlled Phase 3 portion of the trial were the determination of efficacy, safety and, in a subset of participants, immunogenicity of CoVLP+AS03. The trial involved 85 sites in Argentina, Brazil, Canada, Mexico, the United Kingdom, and the United States of America. Once at least 160 laboratory-confirmed COVID-19 cases (≥7 days post second vaccination) were collected, and a median safety follow-up of at least 2 months (post second dose) was achieved in at least 3,000 participants in each of the CoVLP+AS03 and placebo groups, the database was cleaned, a snapshot was taken, and the primary efficacy results were calculated. The cut-off dates for inclusion in the vaccine efficacy analyses and safety analyses were August 20^th^, 2021 and October 25^th^, 2021, respectively.

Medicago Inc. was responsible for the overall trial design and oversight, as well as the manufacture of CoVLP. GSK was responsible for the manufacture of AS03 and providing input into trial design. Site selection and monitoring and conduct of the trial was delegated by Medicago to Syneos (Canada). Data were collected by site investigators (complete list of investigators provided in supplementary materials). Data were centralized and analyzed in conjunction with Syneos. PCR testing and genetic analysis of viral samples was performed by Viroclinics-DDL (Netherlands).

Safety oversight was provided by the Safety Monitoring Team (SMT) that reviewed safety data on a regular basis in a blinded manner. This included data on AESIs including potential Immune Mediated Disorders (pIMDs), Anaphylaxis and severe allergic reactions, Vaccine-associated enhanced diseases (VAED), or Vaccine-associated enhanced respiratory diseases (VAERD), with triggers in place to escalate to the IDMC if a potential safety signal was identified. In parallel an unblinded medical monitor was reviewing unblinded data on a real time basis to escalate to the IDMC if a stopping rule was met. The safety and efficacy data were also reviewed by the IDMC, to confirm that the primary efficacy endpoint had been met, and that the benefit/risk profile was positive.

The trial is ongoing and, at the time of writing, the investigators remained unaware of participant-level treatment assignments. Limited team members have unblinded access to the data to facilitate submission of clinical safety findings to regulatory agencies and the IDMC. All other trial staff and participants remain unaware of treatment assignments.

### Participants and Randomization

Study participants were adults aged 18 years or older and included younger adults aged 18-64 (Population 1), older adults aged 65 or more (Population 2) and adults aged 18 or more with significant comorbidities (Population 3). High-risk comorbidities in Population 3 included, but were not limited to obesity, hypertension, diabetes (type 1 or 2), chronic obstructive pulmonary disease, cardiovascular disease, asthma and immunocompromise due to treatment-controlled HIV, organ transplant, or receipt of cancer chemotherapy (full details in Supplementary Table S2). Participants had not previously received any SARS-CoV-2 vaccine and must not have had a history of virologically confirmed COVID-19. Full inclusion and exclusion criteria are detailed in the trial protocol (available in supplementary materials).

Participants were assigned in a 1:1 ratio to receive either CoVLP+AS03 or placebo. The vaccine candidate or placebo (0.5 mL) were injected intramuscularly, 21 days apart, in contralateral (when possible) deltoid muscles by an unblinded site staff member. Participants were observed by blinded site staff responsible for safety evaluations for 30 minutes after each vaccination.

### Trial Vaccine

The CoVLP vaccine candidate, previously described in detail ^42^, is composed of full-length spike (S) protein in a pre-fusion stabilized configuration from SARS-CoV-2 (strain hCoV-19/USA/CA2/2020) incorporating the modifications: R667G, R668S, R670S, K971P, and V972P expressed in the leaves of *Nicotiana benthamiana* by *Agrobacterium*-based transient transfection. High-level expression of the S protein results in formation of S trimers at the plasma membrane followed by spontaneous budding of CoVLPs. AS03 adjuvant is an oil-in-water emulsion containing DL-α-tocopherol and squalene, supplied by GSK. Immediately prior to use, 2.5 mL of CoVLP and 2.5 mL of AS03 are mixed in a multidose vial to obtain 10 vaccine doses of 0.5 mL each. Each dose of the vaccine contained 3.75 μg of CoVLP formulated in phosphate-buffered saline (PBS) with polysorbate 80, 11.86 mg of DL-α-tocopherol and 10.69 mg of squalene. The placebo was composed of 0.5 mL PBS with polysorbate 80.

### Safety Assessments

The safety analysis included all data collected as of October 25^th^, 2021. The safety analysis set (SAS) used to evaluate unsolicited data including AESIs and SAEs comprised 24,076 participants: 12,036 in the CoVLP+AS03 arm, and 12,040 in the placebo arm. Reactogenicity data for solicited AEs were analysed for a subset (N=4,136 in vaccine arm and N=3,683 for placebo) of participants who had received both doses following the protocol-prescribed dosing regimen and who had completed at least 2 months of safety follow-up, post dose 2.

Solicited local or systemic adverse events within 7 days of receiving each dose were collected using paper or electronic diaries. Unsolicited AEs were monitored for 21 days after each dose, while SAEs, MAAEs, AEs leading to withdrawal, AESIs (including VAED/VAERD, anaphylaxis and severe allergic reactions, pIMDs), and deaths are being monitored throughout the study. AE grading criteria as well as pre-defined study specific stopping rules for safety reasons are detailed in the clinical study protocol (available in supplementary materials).

### Efficacy Assessments

The primary efficacy endpoint was the prevention of symptomatic SARS-CoV-2 infection seven or more days after receipt of the second dose (Primary Vaccine Efficacy; PVE). COVID-19 cases were adjudicated by a subcommittee of the IDMC, blind to group assignment, and were defined by the presence of at least one of the following symptoms: fever or chills, cough, shortness of breath or difficulty breathing, fatigue, muscle or body aches, headache, loss of taste or smell, sore throat, congestion or runny nose, nausea or vomiting, diarrhea, and a positive SARS-CoV-2 test (on nasopharyngeal (NP) or nasal swabs) by quantitative reverse-transcriptase polymerase chain reaction (qRT-PCR) performed by a central virological laboratory (ViroClinics-DDL: Rotterdam, Netherlands) that provided both qualitative (i.e.: positive-negative) and quantitative results (i.e.: viral load: copies/mL; cp/ml).

Efficacy assessments that can be reported at this time are vaccine efficacy in preventing laboratory-confirmed moderate and severe COVID-19, viral load at the time of diagnosis, and efficacy of the vaccine in preventing laboratory-confirmed COVID-19 by variants seven days or more after the second dose. COVID-19 severity assessment was based on FDA guidance criteria and severe COVID-19 was further defined in the protocol.

Viral load was determined by Viroclinics using RT-qPCR targeting the nucleocapsid gene. Primers and assay conditions were based on CDC N1 assay ^44^. The resulting cycle threshold (Ct) values were plotted against a four-point standard curve with a known number of log10 cp/mL run together with the real-time PCR. Ct values were converted to log10 cp/mL based on the slope and intercept of the standard curve for each analysis.

### Viral Sequencing

Sequencing of viral genomes from swabs was performed by Viroclinics-DDL. Total nucleic acid was extracted from samples using MagMAX™ Total Nucleic Acid Isolation Kits (ThermoFisher) and full-length amplification of the target RNA, i.e., RNA encoding S, was performed by nested RT-PCR. Next-generation sequencing platforms (MiSeq and NextSeq Illumina) were deployed to analyze the sequences of the amplicons and the SARS-CoV-2 Wuhan-Hu-1 (GenBank: MN908947.3) was used as the reference sequence for amplicon mapping. A detailed protocol is included in the supplementary material section.

### Statistical Analysis

The full statistical analysis plan is included as a supplementary material.

Safety analyses were descriptive in nature and summarized as counts and percentages. No statistical tests were performed. Safety analyses presented herein include solicited and unsolicited adverse events occurring on or after vaccination and coded as per Medical Dictionary for Regulatory Activities (MedDRA) v24.0.

Efficacy analyses of the primary endpoint included all laboratory confirmed cases that met the definition of a COVID-19 case as adjudicated by the IDMC subcommittee. In order for a COVID-19 case to be considered as a primary endpoint, symptoms had to start 7 or more days after second dose and before the subject was unblinded to study treatment or was administered a currently deployed COVID-19 vaccine. Symptom start was required to occur on or before

August 20^th^, 2021, to be included in the analysis of the primary endpoint. The IDMC subcommittee provided confirmation that a COVID-19 case met the requirements for the primary endpoint and assessed severity according to FDA criteria ^45^.

Vaccine efficacy was calculated as 100 × (1 – incidence rate ratio) where the incidence rate ratio is defined as the ratio of person-years rate of COVID-19 cases in the CoVLP+AS03 group relative to the COVID-19 cases in the placebo group. For both analysis sets, censoring was performed at the earlier of any of the following events, namely, when the subject experienced their first virologically confirmed COVID-19 case (date of first symptoms), the study database was frozen for the primary analysis, the date subject was unblinded, the date subject received an approved or authorized COVID-19 vaccine, or the date of subject completion/withdrawal from the study. The VE success criterion for the primary efficacy endpoint was defined as a ≥50 %-point estimate and a >30 % lower limit of the 95% CI. Assuming the number of cases in each arm followed a Poisson distribution, then conditioning on the total number of cases and the ratio of person-time of follow up, the exact 95% CIs for the incidence rate ratio (IRR), and hence for VE = 1-IRR, was obtained assuming a binomial distribution with mid-P adjustment ^46^. Vaccine efficacy for the secondary endpoints was analyzed using the same methods as for the primary endpoint, except for VE success criterion being defined as >0% lower limit of the 95% CI. Cumulative incidence curves were calculated using the Kaplan-Meier method.

Median differences in viral load values measured as log10 copies/mL were compared between subjects who received CoVLP+AS03 or placebo by strain and overall using the Wilcoxon rank-sum test. Viral load values under the detection limit (log10 2.08) were set to half of that limit.

## Results

### Context in which the study was performed

Unlike many earlier Phase 3 efficacy studies, the current trial was conducted at a time of actual or anticipated roll-out of one or more emergency-use SARS-CoV-2 vaccines. Although the pace and penetration of these roll-out campaigns varied widely across countries and regions, the recruitment of elderly volunteers and those with high-risk comorbidities became progressively more difficult over time, leading to much smaller numbers of participants among healthy older adults and adults with comorbidities than originally planned. Furthermore, as countries/regions enlarged the populations eligible for vaccination, a steadily growing number of enrolled participants in all study populations chose to exercise their Protocol-sanctioned option to be unblinded in order to access a deployed vaccine. This led to a progressive loss of participants in both arms of the study through early withdrawal and unblinding with a growing imbalance between the CoVLP+AS03 and placebo arms of the study that was managed by using person-year denominators to calculate all efficacy outcomes, thus taking into account the accrued amount of follow-up for each participant in the study. These impacts were anticipated in the study design that sought to capture cases as quickly as possible to minimize delays in providing vaccines to placebo recipients and mitigate the risk of withdrawals/unblinding events. Among the important design elements was the use of a single COVID-19-compatible symptom to trigger PCR testing that likely led to early identification of cases as well as the detection of a high proportion of minimally symptomatic cases. Finally, unlike many of the early randomized-controlled trials (RCT) of candidate SARS-CoV-2 vaccines that were confronted with either the ancestral (Wuhan) strain only or a limited spectrum of variants, the current study was performed at a time of active viral evolution and the circulation of multiple variants with increased transmissibility ^47^, resistance to vaccine-induced immunity or both (see Supplementary Figure 1).

### Demographic and baseline clinical characteristics

Participants ≥18 years of age were tested for SARS-CoV-2 antibodies at 85 sites in the USA (N=41), Canada (N=14), the United Kingdom (N=11), Brazil (N=7), Mexico (N=7), and Argentina (N=5), using a commercial ELISA that targets the nucleocapsid (N) protein (ElecSys, Roche Diagnostics). Provided the participant had no previous history of virologically confirmed COVID-19, both seronegative and seropositive participants were enrolled.

Participant disposition and flow diagram are presented in Figure 1. Participant demographics in the PP set are presented in Table 1 and participant demographics in the ITT set are presented in Supplementary Table S1. Of the 25,170 individuals recruited, 24,141 were randomized (ITT set) and 24,076 received one or more study injections. Of these, 20,090 participants received two full vaccinations as scheduled (PP set): 18,150 healthy adults 18-64 years of age, 109 healthy older adults aged 65 years or more (0.4%), and 1,831 adults ≥18 years of age with high-risk comorbidities (7.5%) (listed in Supplementary Table S2). In the PP set, 10,060 were male (50.1%) and 10,030 were female (49.9%). The racial distribution, in order of decreasing frequency, was 88.8% White or Caucasian, 7.0% Black or African American, 1.2% Asian, 0.2% American Indian or Alaska Native, and <0.1% Native Hawaiian or Other Pacific Islander. Two percent reported multiple races and <0.1% reported other or had missing racial data. Overall, 83.3% of participants reported Hispanic or Latinx ethnicity, 16.3% were non-Hispanic or Latinx and 0.4% did not report their ethnicity. The median age was 29. The youngest participant was 18 and the oldest was 86.

**Figure 1.**
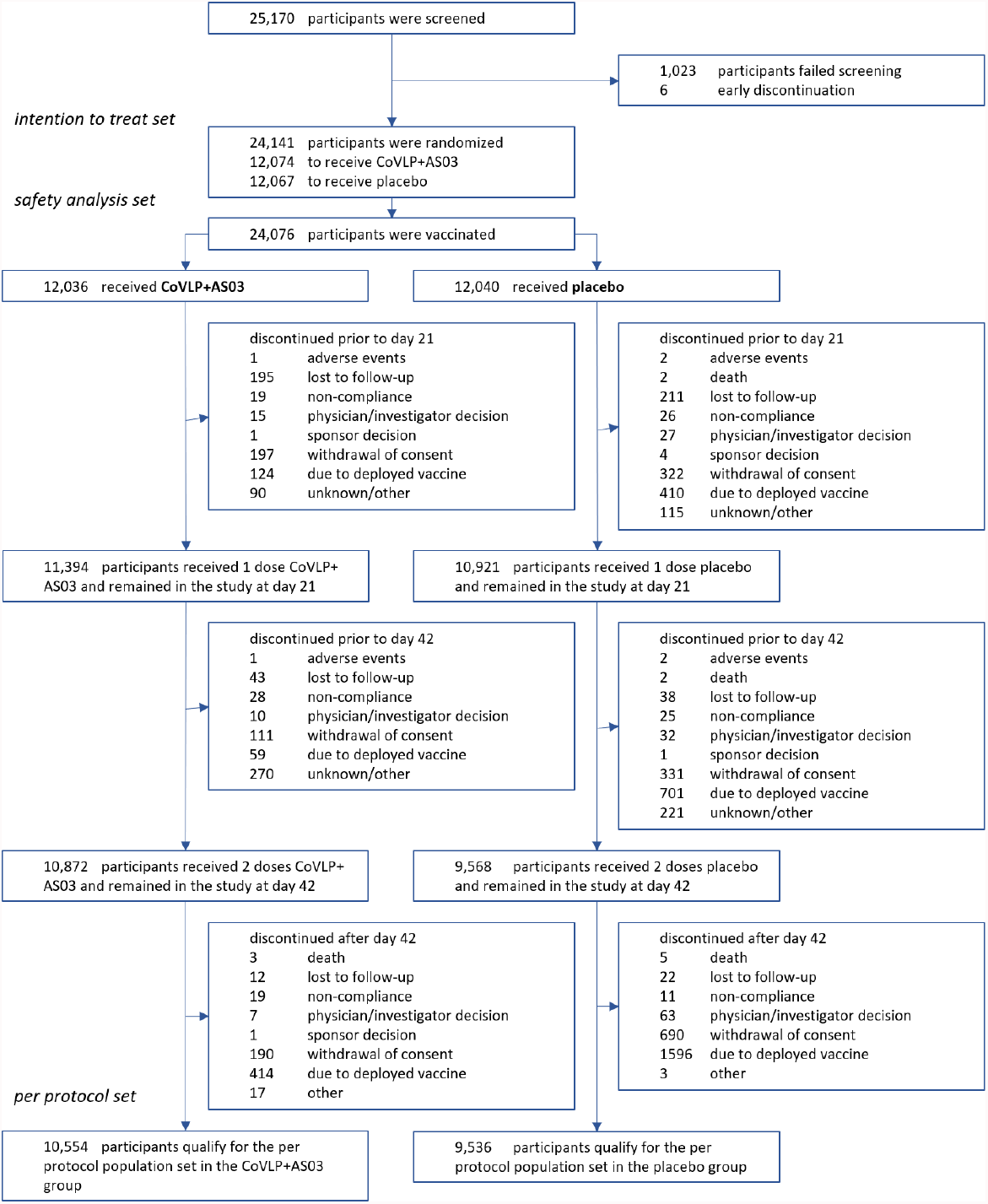
CONSORT Flow Diagram.

**Table 1:** Summary of Demographics in the Per Protocol Set (NCT04636697) ‘N’ is the number of participants in a population, while ‘n’ represents the number of participants in a category. Race and/or ethnic group were reported by the participants, who could have listed more than one category. SD: Standard deviation; Min.: Minimum; Max.: Maximum.

|  | Healthy Adults |  | Older Adults (65+) |  | Adults with Comorbidities |  | All Participants |  |
| --- | --- | --- | --- | --- | --- | --- | --- | --- |
|  | CoVLP + AS03 | Placebo | CoVLP + AS03 | Placebo | CoVLP + AS03 | Placebo | CoVLP + AS03 | Placebo |
| **Participants (N)** | 9,541 | 8,609 | 55 | 54 | 958 | 873 | 10,554 | 9,536 |
| **Sex, n (%)** |  |  |  |  |  |  |  |  |
| Male | 4,764 (49.9) | 4,324 (50.2) | 30 (54.5) | 27 (50.0) | 474 (49.5) | 441 (50.5) | 5,268 (49.9) | 4,792 (50.3) |
| Female | 4,777 (50.1) | 4,285 (49.8) | 25 (45.5) | 27 (50.0) | 484 (50.5) | 432 (49.5) | 5,286 (50.1) | 4,744 (49.7) |
| **Race, n (%)** |  |  |  |  |  |  |  |  |
| American Indian/ Alaska Native | 21 (0.2) | 15 (0.2) | 0 | 0 | 1 (0.1) | 4 (0.5) | 22 (0.2) | 19 (0.2) |
| Asian | 116 (1.2) | 94 (1.1) | 1 (1.8) | 0 | 9 (0.9) | 7 (0.8) | 126 (1.2) | 101 (1.1) |
| Black/ African American | 449 (4.7) | 434 (5.0) | 7 (12.7) | 9 (16.7) | 101 (10.5) | 97 (11.1) | 557 (5.3) | 540 (5.7) |
| Multiple | 177 (1.9) | 183 (2.1) | 1 (1.8) | 0 | 22 (2.3) | 17 (1.9) | 200 (1.9) | 200 (2.1) |
| Native Hawaiian or Other Pacific Islander | 19 (0.2) | 14 (0.2) | 0 | 0 | 2 (0.2) | 4 (0.5) | 21 (0.2) | 18 (0.2) |
| White or Caucasian | 8,720 (91.4) | 7,835 (91.0) | 45 (81.8) | 45 (83.3) | 817 (85.3) | 743 (85.1) | 9,582 (90.8) | 8,623 (90.4) |
| Other | 1 (<0.1) | 1 (<0.1) | 0 | 0 | 0 | 0 | 1 (<0.1) | 1 (<0.1) |
| Not Reported | 35 (0.4) | 29 (0.3) | 0 | 0 | 5 (0.5) | 1 (0.1) | 40 (0.4) | 30 (0.3) |
| **Ethnicity, n (%)** |  |  |  |  |  |  |  |  |
| Hispanic or Latinx | 8,076 (84.6) | 7,171 (83.3) | 13 (23.6) | 22 (40.7) | 761 (79.4) | 698 (80.0) | 8,850 (83.9) | 7,891 (82.7) |
| Not Hispanic or Latinx | 1,428 (15.0) | 1,409 (16.4) | 42 (76.4) | 32 (59.3) | 196 (20.5) | 171 (19.6) | 1,666 (15.8) | 1,612 (16.9) |
| Not Reported/ Specified | 37 (0.4) | 29 (0.3) | 0 | 0 | 1 (0.1) | 4 (0.5) | 38 (0.4) | 33 (0.3) |
| **Age at consent (years)** |  |  |  |  |  |  |  |  |
| Mean (SD) | 31.8 (10.98) | 31.9 (11.13) | 69.9 (4.30) | 69.8 (5.09) | 38.7 (13.74) | 40.2 (14.37) | 32.6 (11.72) | 32.9 (12.01) |
| Median | 29.0 | 29.0 | 69.0 | 68.0 | 37.0 | 39.0 | 29.0 | 29.0 |
| Min, Max | 18, 64 | 18, 64 | 65, 80 | 65, 86 | 18, 82 | 18, 86 | 18, 82 | 18, 86 |

### Efficacy

Up to the cut-off date of August 20^th^, 2021, a total of 401 possible COVID-19 cases were identified in the study population. Of these, based on time of occurrence relative to vaccination, 176 were predicted to potentially contribute to determination of the PVE endpoint.

Adjudication confirmed the applicability of 165 of these cases (10 were removed due to unblinding prior to diagnosis of COVID-19 and 1 case was determined not to have met the PVE criteria). The PVE analysis was determined based on these 165 adjudicated cases (ITT set; 157 in the PP set).

Among the 20,090 participants in the PP set, 118 in the placebo group (9,536 participants) and 39 in the CoVLP+AS03 group (10,554 participants) developed COVID-19 ≥7 days after receiving the second dose. The incidence rate in the placebo group was 0.179 per 1,000 person-years (95% CI: 0.150, 0.215) and was 0.052 in the CoVLP+AS03 group (95% CI: 0.038, 0.071). This corresponds to an overall vaccine efficacy of 71.0% (95% CI: 58.7, 80.0, see Figure 2) irrespective of Day 0 serostatus. In the ITT set of 24,141 participants, 125 in the placebo group (12,067 participants) and 40 in the CoVLP+AS03 group (12,074 participants) developed COVID-19 ≥7 days after receiving the second dose. This corresponds to a vaccine efficacy of 69.5% (95% CI: 56.7, 78.8; see Figure 2 for incidence rates). VE success for the primary efficacy endpoint was defined as a ≥50 % point estimate and a lower limit of the 95% CI >30%. These primary outcome criteria were met in both the ITT and PP sets.

**Figure 2.**
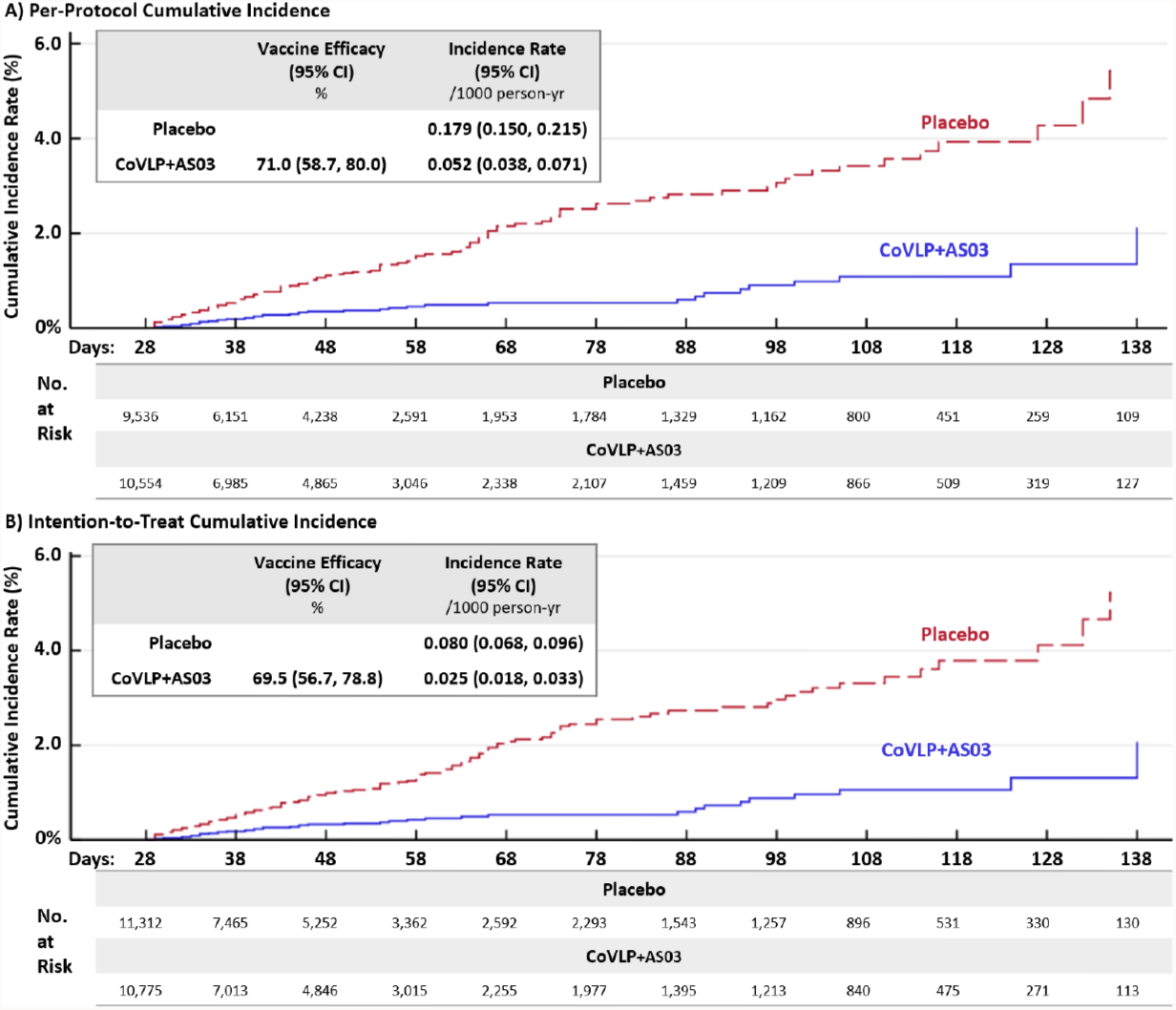
Cumulative Incidence Curves from Seven Days Post Second Injection.

As shown in Figure 3 for the PP set and in Supplementary Table S3 for the ITT set, VE of CoVLP+AS03 was also separately evaluated in healthy participants 18-64 years of age, healthy participants ≥ 65 years of age, and participants ≥ 18 years of age with significant comorbidities. The VE point estimates in the PP set were 70.9% (95% CI: 57.7, 80.4) and 76.8% (95% CI: 21.5, 94.8) for healthy adults aged 64 or less and in adults with significant comorbidities, respectively, and 68.9% (95% CI: 55.0, 78.9) and 78.7% (95% CI: 30.2, 95.1) in these populations in the ITT, respectively irrespective of baseline (Day 0) serostatus. There were only two cases of COVID-19 in healthy older adults aged 65 or more, one in the placebo group and one in the CoVLP+AS03 group, precluding accurate assessment of VE in that population.

**Figure 3.**
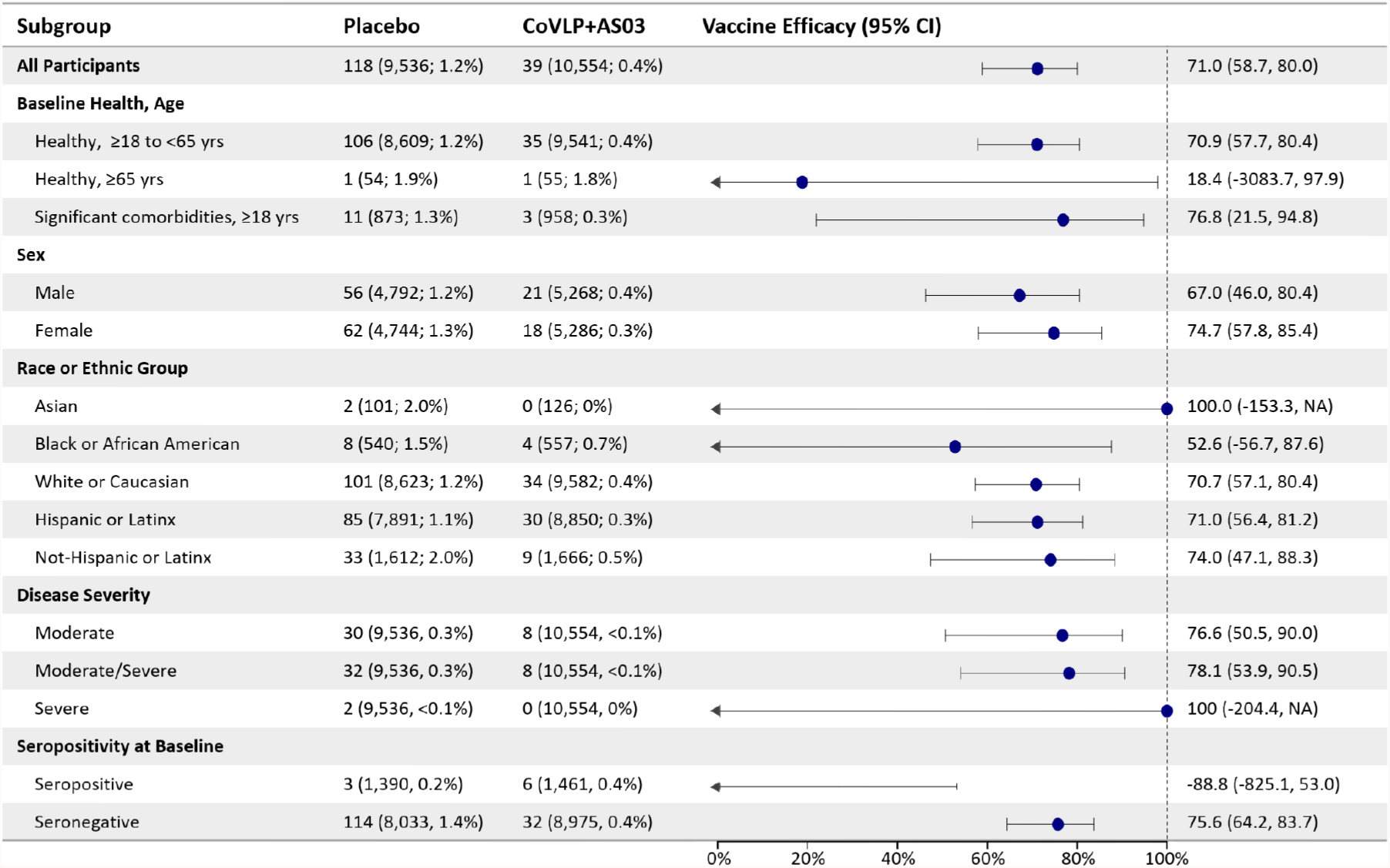
Efficacy by Subgroup.

Vaccine efficacy in preventing moderate (*ad hoc* analysis) or severe disease was determined to be 78.1% (95% CI: 53.9, 90.5) in the PP population and 78.8% (95% CI: 55.8, 90.8) in the ITT set. Among subjects who were seronegative at Day 0, the overall VE against moderate-to-severe disease was 84.5% (95% CI: 62.0,94.7) in the PP set and 86.0% (95%CI: 66.2, 95.1) in the ITT set. There were only 3 severe cases of COVID-19 in the 165 cases used for the PVE analysis (2 hospitalized) and all were in the placebo group. The VE point estimates were determined by gender, race, and baseline (Day 0) serostatus and are presented in Figure 3 (PP set) and in Supplementary Table S3 (ITT set).

### Variant-Specific Efficacy

Supplementary Figure 1 illustrates the incidence of COVID-19 by variant in the participating countries (colored bars, left Y-axis) as well as the number of participants with COVID-19 contributing to the PVE analysis (red lines, right Y-axis). Up to the cut-off date of August 20^th^, 2021, COVID-19 cases included in the PVE analysis were primarily identified in Argentina (n=59), Brazil (n=53), and the United States of America (n=47), and with much smaller numbers in the United Kingdom (n=4) and Canada (n=2).

Based on genomic sequences shared via GISAID, the global data science initiative ^48^, the primary variants circulating during the study varied widely by country (see Supplemental Figure 1). The Delta and Gamma strains were the dominant strains in Argentina and Brazil with lesser contributions from Alpha and Lambda variants while Alpha and Delta variants dominated in North America and the UK with lesser contribution from the Gamma variant in Canada and the USA.

Of the 157 cases included in the per protocol PVE analysis, sequence data are available for 114 (72.6%) and a further 21 (13.4%) could not be sequenced due to very low viral copy numbers in the samples. All sequenced strains were either variants of concern (VoC) or variants of interest (VoI). No case included in the PVE analysis was caused by an ancestral (Wuhan) strain virus. The most frequently sequenced variants were Delta (50, 43.9%) and Gamma (52, 45.6%) with fewer numbers of Alpha (5, 4.4%), Mu (4, 3.5%) and Lambda (3, 2.6%) variants. There were no cases caused by Beta or Omicron variants among those infected.

As shown in Figure 4, for the PP set, overall vaccine efficacy point estimates were 75.3% (95% CI: 52.8, 87.9) and 88.6% (95% CI: 74.6, 95.6) for the dominant Delta and Gamma variants, respectively, and were 100% with much broader 95% confidence intervals for the small numbers of Alpha (95% CI: 28.0, -na-), Lambda (95% CI: -50.3, -na-), and Mu variants (95% CI: 2.3, -na-). The corresponding point estimates for the ITT set ranged from 74.0% and 87.8% for Delta and Gamma variants respectively to 100% for the other three variants (see Supplemental Table 3).

**Figure 4.**
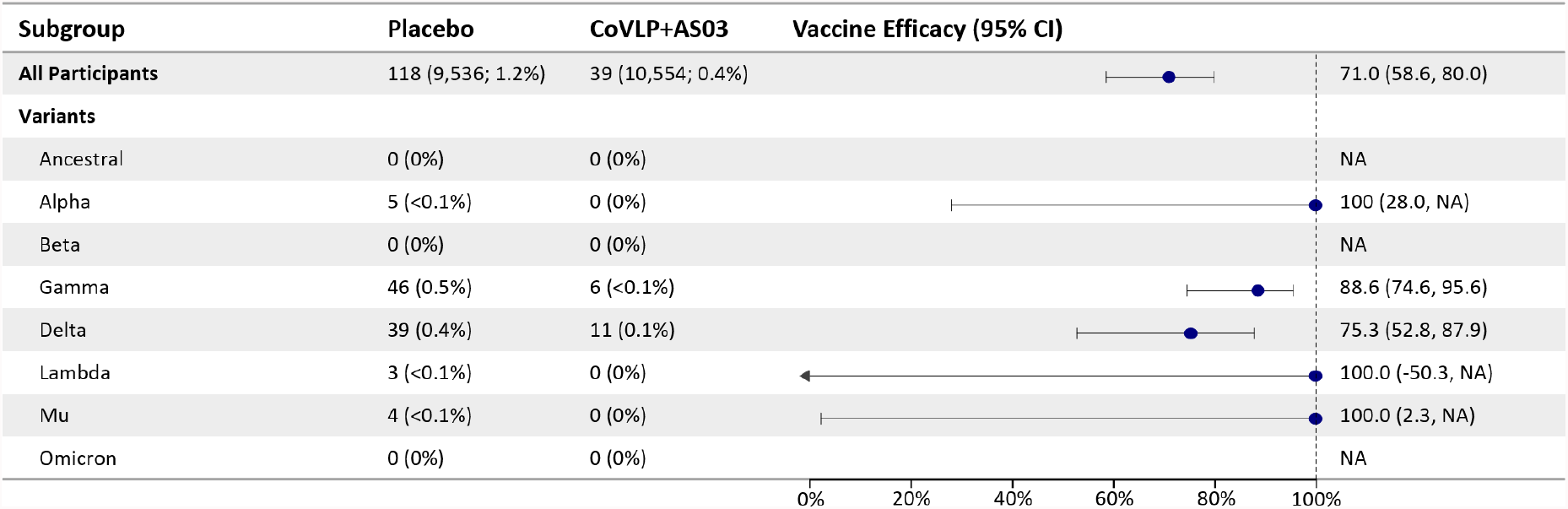
Efficacy by Circulating Variant.

Sequencing success was markedly different between the CoVLP+AS03 (43.6%) and Placebo groups (82.0%) in the PP set (34.6% versus 83.2%, respectively in the ITT set). Analysis of viral loads at the time of diagnosis revealed a >100-fold difference overall between the CoVLP+AS03 and placebo cases (log10 3.46 versus log10 5.65 copies/mL, respectively; see Figure 5). Among the PP set, cases for which sequencing failed [i.e.: PCR-positive but Sequence-negative (PCR^+^/Seq^-^)], 14 were in the CoVLP+AS03 group and 7 were in the placebo group. For these PCR^+^/Seq^-^ cases, the median viral load was at or below the limit of detection of the assay used (120 copies/mL) while the median viral load for the cases from which sequence information could be obtained was >500,000 copies/mL. Viral loads in the breakthrough Delta and Gamma cases were 42-fold and 269-fold lower in the CoVLP+AS03 group compared to the placebo group (log10 3.65 versus log10 5.27 and log10 3.78 versus log10 6.21, respectively; See Figure 5). A similar trend of markedly lower viral loads was observed in breakthrough cases classified as either mild (log10 3.56 versus log10 5.70 or 138-fold) or moderate (log10 2.85 versus log10 5.48 or 426-fold) in the CoVLP+AS03 and placebo groups, respectively.

**Figure 5.**
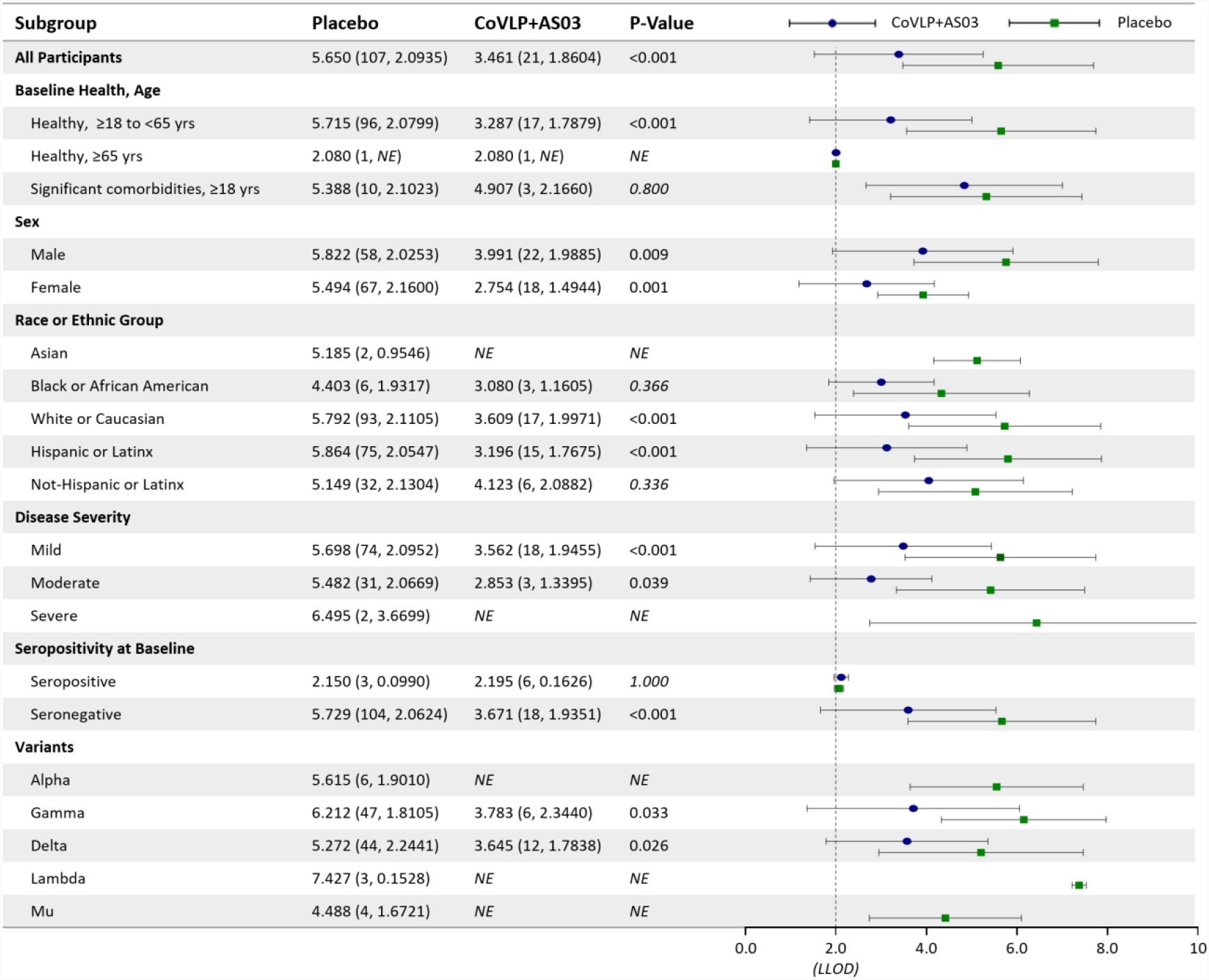
Viral Load.

### Tolerability and Safety

Solicited AEs up to 7 days after first or second injection for both CoVLP+AS03 and placebo recipients were collected and analyzed for 7,819 participants (4,136 CoVLP+AS03 recipients and 3,683 placebo recipients) and are shown in Figure 6. Overall, both solicited local and systemic AEs were predominantly mild to moderate in intensity and transient in nature, lasting 1-3 days on average.

**Figure 6.**
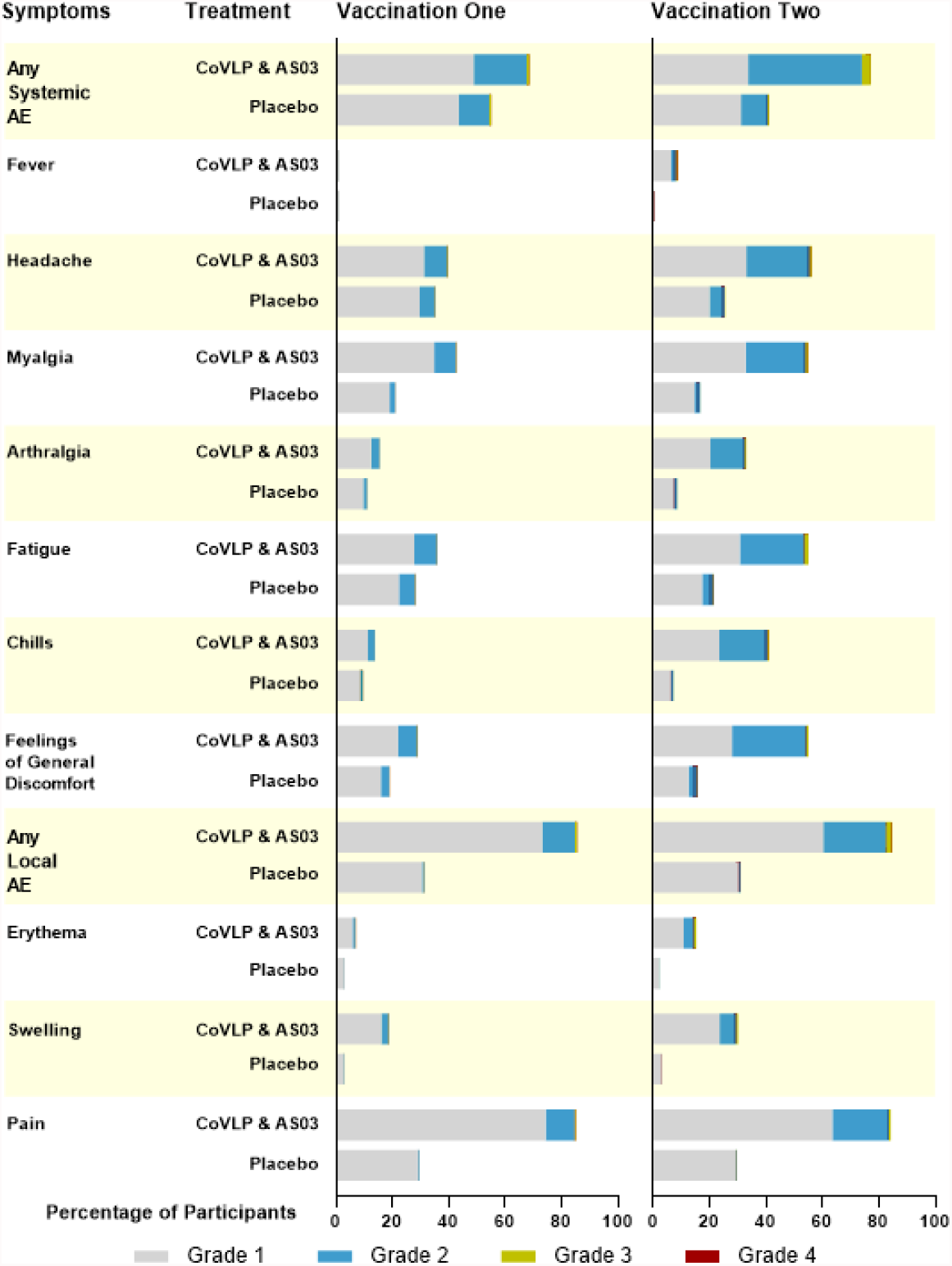
Solicited Adverse Events.

As shown in Supplementary Table S4, more participants who received the CoVLP+AS03 (3,819; 92.3%) than those who received placebo (1,677; 45.5 %) reported local solicited AEs after receiving first and/or second treatment; driven largely by pain at injection site. None of the solicited local AEs were potentially life-threatening (Grade 4). The incidence of Grade 2 and Grade 3 local AEs was higher after the second vaccination. Severe (Grade 3) solicited local AEs were reported by 33 (0.8%) and 85 (2.1%) after the first and second injections respectively among those who received CoVLP+AS03, and 2 (<0.1%) and 1 (<0.1%) respectively among the participants who received placebo.

As shown in Supplementary Table S5, similar to local solicited AEs, solicited systemic AEs were also more common in the participants who received CoVLP+AS03 (3,612; 87.3%) compared to those who received placebo (2,394; 65.0%) after receiving first and/or second treatment. Both the frequencies and intensities of these events increased after the second dose compared to the first (Supplementary Table S5). The more common systemic AEs in both groups were headache, fatigue, myalgia, and a general feeling of discomfort. Fever was reported in 1.1% after the first dose and was reported by 8.6% after the second dose in CoVLP+AS03 recipients. There were 42 participants (1.0%) with Grade 3 solicited systemic AEs after the first injection and 129 (3.1%) after the second injection in the CoVLP+AS03 group, and 24 (0.7%) and 20 (0.5%) in the placebo group after the first and second injections respectively. Three participants reported Grade 4 systemic solicited AEs, 2 (<0.1%) in the CoVLP+AS03 group and 1 (<0.1%) in the placebo group; all occurred after the second injection.

For the analysis of the unsolicited AEs, safety data available from all participants in the SAS were included. The study measured the occurrence, intensity, and relationship of unsolicited AEs for 21 days after each dose, while SAEs, MAAEs, AEs leading to withdrawal, AESIs (including VAED/VAERD, anaphylaxis and severe allergic reactions, pIMDs), and deaths were monitored for 21 days after each dose (Supplementary Table S6) and then from day 43 to day 201 (Supplementary Table S7). The incidence of unsolicited AEs after receiving first and/or second treatment was slightly higher in CoVLP+AS03 recipients than in placebo recipients (22.7% vs 20.4% during the 21 days after vaccination, and 4.2% vs 4.0% during the 43 to 201 days after vaccination). Unsolicited preferred term (PT) events with a frequency ≥1% after receiving first and/or second vaccination are presented in Supplementary Table S8.

The frequency of SAEs reported in subjects was similar between vaccine (24: 0.2%) and placebo recipients (16: 0.1%) up to 21 days after receiving first and/or second treatment. Between Day 43 and Day 201, SAEs were reported by 19 (0.2%) and 22 (0.2%) of the participants in the CoVLP+AS03 and placebo groups, respectively after receiving first and/or second treatment. No SAE was assessed as related to the investigational product in CoVLP+AS03 group and one subject in the placebo group reported 2 related SAEs (aortic thrombosis and peripheral artery thrombosis). No significant imbalance or safety concern was noted in MAAEs, AESIs, AEs leading to withdrawal, deaths, or specific AEs reported after vaccination with currently authorized vaccines (Bell’s palsy, myocarditis, thrombotic events, etc. ^49^; see Supplemental Table S9). There were no deaths related to the vaccine in the study.

## Discussion

Despite the challenges of conducting a Phase 3, placebo-controlled vaccine efficacy study in the face of international vaccination roll-out campaigns and rapid viral evolution, CoVLP+AS03 met the primary efficacy endpoint of the study. Two doses of CoVLP+AS03 delivered 21 days apart provide substantial protection against symptomatic COVID-19 caused by a range of SARS-CoV-2 variants including Alpha, Delta, Gamma, Lambda, and Mu.

The vaccine provided >70% protection against symptomatic COVID-19 of any severity in a diverse adult population including those with high-risk comorbidities irrespective of baseline serostatus. While vaccine efficacy in healthy adults over the age of 65 could not be determined due to ethical considerations that limited the number of older adults in the study, two doses of CoVLP+AS03 were previously shown to induce a neutralizing antibody response in healthy older adults that is indistinguishable from that seen in healthy adults ^41^, suggesting that CoVLP+AS03 may provide comparable protection across the adult age range.

Prevention of severe disease and hospitalization, both to improve health outcomes and to alleviate health care resource constraints, remains a critical objective of national vaccination campaigns. While there were only two PP cases of severe disease in the trial and three in the ITT set (two of which required hospitalization), all were in the placebo group. Furthermore, in an *ad hoc* analysis, overall vaccine efficacy against combined moderate-severe disease was 78.1%. Since the concentration of virus in the upper respiratory tract is a major determinant of sequencing success, the fact that the infecting strain could be identified in only 43.6% of the CoVLP+AS03 cases compared to 82% of the Placebo cases suggested that the viral loads in the PCR^+^Seq^-^ ‘breakthrough’ cases in the vaccinated group would be low and that these cases would be mild, both of which proved to be true. Although completely asymptomatic individuals can have high viral loads in their upper respiratory tracts and there are conflicting data on the relationship between viral load at diagnosis and disease progression/severity, many groups have demonstrated that viral load in the upper respiratory tract can be a predictor of disease severity in COVID-19 patients ^50-53^. All of the PCR^+^Seq^-^ cases in current study were adjudicated to be mild and interestingly, the viral loads in these cases in either group were at or near the limit of detection of the RT-qPCR assay used (log10 2.08 copies/mL) with a mean value ∼3715x lower than the PCR^+^Seq^+^ cases in the placebo group (log10 5.65 copies/mL: Figure 5). These observations suggest that the overall VE point estimate for CoVLP+AS03 was, at least to some extent, drawn down by mild, PCR^+^Seq^-^ breakthrough cases with very low viral loads. Indeed, the VE point estimate for the very mild, PCR^+^Seq^-^ cases was -75.3% (95% CI: -364.0, 28.6) which explains the difference between the overall VE estimate (71%) and both the strain-specific estimates (75.3-100%) and the estimate for the prevention of moderate or severe disease (78.1%). A large real-world evidence (RWE) study in the UK recently demonstrated the dilutive effect of low viral load cases on VE estimates for both mRNA and Adenovirus-vectored vaccines ^54^. During the period when the Delta variant predominated, for example, the efficacy of an mRNA vaccine for high viral load disease was 86% (Ct values <30) but fell to 71% for low viral load illness (Ct values ≥30). A unique feature of the current study, which helped identify cases as quickly as possible in the rapidly evolving pandemic, was the use of a single COVID-19-compatible symptom to trigger PCR testing. While this undoubtedly led to faster case accrual, it is likely that the use of a single symptom trigger may have led to the inclusion of a disproportionate number of very mild cases, and consequently to an underestimation of overall PVE relative to studies that used more restrictive clinical criteria to trigger swab collection.

Analysis of viral load over time after diagnosis was included in this study as a secondary outcome to look at the impact of vaccination on the magnitude and duration of viral shedding. However, an initial observation of a 2-fold difference in sequencing success between the CoVLP+AS03 and placebo cases suggested there might be important differences in the viral load at diagnosis and prompted an immediate analysis of these results. To our knowledge, this is the first Phase 3 RCT that has considered viral load as a parameter in the characterization of cases. This analysis in different subgroups revealed that breakthrough PCR^+^/Seq^-^ cases in the CoVLP+AS03 group had between 42-and 420-fold less virus in the nasal passages at the time of diagnosis compared to the placebo group. Although this represents only a single time-point in the evolution of each case, follow-up swabs were collected every other day from these subjects while symptoms persisted to address one of the study’s secondary outcomes (i.e.: duration and magnitude of viral shedding). Analysis of these swabs is underway but, given the striking initial differences in viral load at the time of diagnosis, consistent with recent analysis examining viral load in Adenoviral vector-and mRNA-vaccinated individuals ^55^, it seems likely that vaccination with CoVLP+AS03 had significant virologic impact in the breakthrough cases with possible implications for both disease severity and reduced transmission.

While the recently-emerged Omicron variant threatens to spread rapidly, the dominant variants at the time this study was conducted were Alpha and Delta in North America and the UK, and Gamma and Delta in South America (Supplementary Figure 1) with lesser contributions from Lambda and Mu variants. Like all other currently deployed vaccines, CoVLP+AS03 was designed to target the ancestral (Wuhan) strain of SARS-CoV-2 but not a single case of COVID-19 caused by this strain was identified in the current study. This trial, along with Clover’s recently announced global study and Novavax’s regional studies, are among the first Phase 3 RCTs to be confronted by a variant-dominant environment ^17,56^. Although vaccines developed during the first waves of the pandemic reported vaccine efficacies as high as 95% ^7,57^, the more recent RCTs and RWE studies performed during successive variant-dominated periods have consistently demonstrated that overall vaccine efficacy is substantially reduced compared to the Wuhan-era, although efficacy against the most severe forms of COVID-19 has generally been preserved. A recent meta-analysis of vaccine efficacy against the Delta variant by platform ^14^ suggested performance of 59% (95% CI: 26.1, 100) for inactivated vaccines, 67.7% (95% CI: 62.3, 72.5) for Adenovirus-vector vaccines, and 77.7% (95% CI: 68.22, 88.59) for mRNA-based vaccines, possibly attributable to increased viral replication and transmissibility of this variant, as well as immune-evasive mutations in the spike protein ^13^.

Although the results from RCTs and RWE studies should only be compared with caution and most RWE studies are influenced by both strain and time post-vaccination ^15,58^, the context in which vaccines are currently ‘asked to perform’ has clearly changed and the overall VE for CoVLP+AS03 of 71% (84.5% in baseline seronegative individuals) with strain-specific VE ranging from 75.3-100% seen in the current study appears to compare favorably to the currently reported effectiveness of other candidates and deployed vaccines ^13-15,17,56,59^. This is particularly true since CoVLP+AS03 was challenged in this study by a range of strains with known immune-evasive mutations, including Delta, Gamma and Mu variants ^60,61^. The challenges faced by CoVLP+AS03 in the on-going Phase 3 study continue with the rapid emergence of the Omicron variant. Active surveillance for cases will continue until the trial is terminated per-Protocol when the placebo recipients are offered vaccination or release from their study commitments and VE of CoVLP+AS03 against this most recent VoC will be calculated.

Overall, CoVLP+AS03 was well-tolerated and the safety profile in the Phase 3 portion of the Phase 2/3 study largely confirmed observations of the smaller Phase 1 study ^42^ and the Phase 2 portion ^41^. Most vaccine recipients reported at least one local or systemic adverse event, the large majority of these events were Grade 1-2 and transient and consistent with past reports of AS03-adjuvanted influenza vaccines ^40^. The safety profile was generally comparable to recently reported solicited and unsolicited safety data for other deployed and candidate SARS-CoV-2 vaccines ^7,57,62,63^. Although 41.8% of participants reported mild-to-moderate and transient chills after the second dose, a documented fever was largely absent after the first dose and only reported by 8.6% of the participants after the second dose. As generally observed for COVID-19 vaccines ^7,57,62,63^, there was an increase in both the frequency and severity of solicited AEs after the second dose compared to the first dose. No safety concerns related to vaccination identified during the study up to the safety data cut-off date of October 25^th^, 2021. Although the number of participants exposed to the CoVLP+AS03 in the clinical development to date remains relatively small (∼13,000 in the Phase 1, 2 and 3 studies) with a relatively short period of follow-up post vaccination, it is nonetheless reassuring that there has been no suggestion of VAED in either a large primate challenge study ^64^ or in the clinical trials ^41,42^. There were no episodes of anaphylaxis or severe allergic reactions reported in the study. It is also reassuring that no imbalance of myocarditis or thrombotic events was observed, and all reported SAEs were considered unrelated to CoVLP+AS03 by the investigators. Although the past 24 months have witnessed an unprecedented growth in vaccine science with the introduction and large-scale deployment of several SARS-CoV-2 vaccines produced using new platforms, this remarkable period of vaccine innovation has not yet run its course. If CoVLP+AS03 is licensed, it will be the first plant-based vaccine approved for human use and one of only a small number of plant-produced biopharmaceuticals ^65^. It may also be the first VLP vaccine for SARS-CoV-2, bringing the potential advantages of this vaccine technology to the fight against COVID-19 ^24^. The plant-based manufacturing platform has a number of natural advantages since it can theoretically be introduced across a wide range of scales from a modular, country-sized unit to a global manufacturing facility. Although the downstream processing and purification procedures are similar across all recombinant protein vaccine platforms, the upstream processes for plant-produced vaccines are based on sunlight and tightly controlled water and growth substrate to support the living plant ‘bioreactor’. As a result, this platform has the potential to promote distributive vaccine (and other biopharmaceutical) production rather than the current highly centralized production model that has contributed to the striking imbalances in the global distribution of SARS-CoV-2 vaccines ^66-68^. Like other VLP vaccines, CoVLP is stable at refrigerator temperatures (2-8°C), making it easier to use in small and remote communities in resource-rich countries as well as in low-and middle-income countries than several of the currently deployed vaccines ^66^. Although much attention has been devoted to the ‘new’ vaccine platforms like the mRNA and Adenovirus-vectored vaccines, there is still clearly a place in the fight against SARS-CoV-2 for more traditional inactivated virion and protein+adjuvant vaccines ^69^. In particular, protein+adjuvant vaccines may be attractive to those who want to be vaccinated but have objecting beliefs or concerns regarding platforms associated with currently available vaccines ^23^. Protein+adjuvant vaccines may also play an important role as an option to boost the responses initiated by other primary vaccination strategies ^59,70^.

Like any Phase 3 study done under pandemic conditions, the current trial had limitations, the most obvious of which are the limited number of older adults and smaller proportion of adults with comorbidities who were able to participate due to on-going vaccine roll-out programs and the relatively short period of follow-up due to the event-driven design. However, immunogenicity data from the Phase 2 portion of the current study demonstrated strong neutralizing antibody and cellular responses in all three study populations after two doses of CoVLP+AS03 ^41^, and 6-month follow-up of CoVLP+AS03 recipients in the preceding Phase 1 study demonstrated that both humoral and cellular responses were durable ^71^. Although relatively few severe cases (three in the ITT set and two in the PP set) and two hospitalizations occurred in this study, all fell into the placebo arm and the overall efficacy against moderate and severe disease combined was 78.1% and 78.8% in the ITT and PP sets, respectively, suggesting that protection against the more severe manifestations of COVID-19 caused by a range of variants was substantial. This study is still ongoing, and a wide range of secondary outcomes will be reported as the data become available.

The primary vaccine efficacy and safety data presented here demonstrate that substantial protection against a range of VoC can be provided by two doses of CoVLP+AS03. New cases continue to be identified in the ongoing Phase 3 study and the efficacy of this new candidate vaccine against the recently-emerged Omicron variant will be determined in the coming months. Overall, the vaccine was generally well-tolerated and no safety signals were detected during the study. Once approved by regulators, this more traditional protein+adjuvant vaccine produced using the novel plant-based platform may be able to make an important contribution to the global struggle against the increasingly complex family of SARS-CoV-2 viruses.

## Author Contributions

All authors contributed significantly to the submitted work. KJH, GH, AIM, AM, JA, JN, ST, MAD, YK, BJW contributed to all aspects of the clinical study from conception to completion. PG, SP, KB, LD, SL, PS, IB, AL, JP, JJW, EP, LT, ASE, LHM contributed to design and execution of the study as well as analysis and presentation of the data. GPM, RSD, EV, FR, RGW, GW, HA, CJ, TB, MAC, MK, FR, FPP contributed to design and oversight of the conduct of the study. All authors contributed to critical review of the data and the writing of the manuscript. All Medicago authors had full access to the data. YK, MAD, BJW made the final decision to submit the manuscript.

## Data Availability

Medicago Inc. is committed to providing access to anonymized data collected during the trial that underlie the results reported in this article, at the end of the clinical trial, which is currently scheduled to be 1 year after the last participant is enrolled, unless granted an extension. Medicago Inc. will collaborate with its partner (GlaxoSmithKline, Wavre, Belgium) on such requests before disclosure. Proposals should be directed to. To gain access, data requestors will need to sign a data access agreement and access will be granted for non-commercial research purposes only.

## Acknowledgements

The authors acknowledge all the volunteers who participated in the study as well as the site investigators and their staff for conducting the studies with a high degree of professionalism. The authors also wish to acknowledge all the employees and contractors of Medicago and GSK who supported the study, directly or indirectly. With regard to supplementary Figure 1, we gratefully acknowledge all data contributors (the authors, their originating laboratories responsible for obtaining the specimens, and the submitting laboratories) for generating the genetic sequence and metadata and sharing it via the GISAID Initiative, on which this figure is based.

## Data Availability

Medicago Inc. is committed to providing access to anonymized data collected during the trial that underlie the results reported in this article, at the end of the clinical trial, which is currently scheduled to be 1 year after the last participant is enrolled, unless granted an extension.

Medicago Inc. will collaborate with its partner (GlaxoSmithKline, Wavre, Belgium) on such requests before disclosure. Proposals should be directed to. To gain access, data requestors will need to sign a data access agreement and access will be granted for non-commercial research purposes only.

## Figure Legends

**Figure 1**

**Trial Profile – Participant Disposition**

Data cut-off for primary efficacy analysis occurred on August 20^th^, 2021. The intention-to-treat (ITT) population was comprised of participants in the randomized population who had no virologic evidence of COVID-19 prior to injection. The per-protocol (PP) population included participants who received two doses and had no major protocol deviations. The safety analysis set (SAS) included all participants who received at least one injection. For details on participant demographics, see Table 1.

**Figure 2**

**Cumulative Incidence of COVID-19 in CoVLP+AS03 Vaccinated and Placebo Control Study Participants**

Cumulative incidence of adjudicated COVID-19 events in the per protocol (a) and intention to treat (b) populations starting 7 days after the second vaccination. Red lines indicate placebo treatment and blue lines indicate CoVLP+AS03 treatment. Vaccine efficacy was calculated as 100 × (1 – incidence rate ratio) where the incidence rate ratio is defined as the ratio of person-years rate of COVID-19 cases in the CoVLP+AS03 group relative to the COVID-19 cases in the placebo group. Events (tick marks) are COVID-19 cases from PCR-positive nasopharyngeal swabs independently confirmed and adjudicated by an IDMC sub-committee.

**Figure 3**

**Subgroup Analysis of Vaccine Efficacy of CoVLP+AS03 to Prevent COVID-19**

Efficacy of CoVLP+AS03 vaccine in preventing COVID-19 in various subgroups within the per protocol population. Subgroup vaccine efficacy was calculated as 100 × (1 – incidence rate ratio) where the incidence rate ratio is defined as the ratio of person-years rate of COVID-19 cases in the CoVLP+AS03 group relative to the COVID-19 cases in the placebo for each subgroup analyzed. Yrs: years; 95% CI: 95% confidence interval; NA: not applicable

**Figure 4**

**Vaccine Efficacy by Variant**

Efficacy of CoVLP+AS03 vaccine in preventing COVID-19 by variant within the per protocol population. Vaccine efficacy by variant was calculated as 100 × (1 – incidence rate ratio) where the incidence rate ratio is defined as the ratio of person-years rate of COVID-19 cases in the CoVLP+AS03 group relative to the COVID-19 cases in the placebo for each variant. Yrs: years; 95% CI: 95% confidence interval; NA: not applicable

**Figure 5 Viral load**

Mean viral loads, presented in log virus copies per mL, are provided for both placebo and CoVLP+AS03 recipients by subgroup. Numbers in parentheses indicate 1) the number of individuals in each group, and 2) standard deviation. To the right, symbols indicate mean log virus copy per mL (blue circles: CoVLP+AS03, green squares: placebo) and horizontal bars indicate standard deviation. P-values as determined by Wilcoxon rank-sum test indicate differences between the placebo and CoVLP+AS03 recipients by subgroup. All analysis is based on the intention to treat (ITT) population set. NE: not estimable.

**Figure 6**

**Solicited Local and Systemic AEs during the 7-Days After the First or Second Doses in both study groups (CoVLP+AS03 vs placebo)**

Participants were monitored for solicited local and systemic Adverse Events (AEs) from the time of vaccination through 7 days after vaccine administration. Participants who reported no AEs or for whom Serious Adverse Events (SAE) data are lacking make up the remainder of the 100% calculation (not shown). For each category, AEs are classified as follows: Grade 1 = Mild; Grade 2 = Moderate; Grade 3 = Severe; Grade 4 = Potentially life threatening. If a participant had the same AE but with different grades, the highest grade was reported. If any of the solicited AEs persisted beyond Day 7 after vaccination, it was recorded as an unsolicited AE. Fever was defined as oral temperature ≥38.0°C.

