## Supplemental Materials for "Efficacy and Safety of a Plant-Based Virus-Like Particle Vaccine for COVID-19 Adjuvanted with AS03"

### Supplementary Appendix

*List of investigators*

| Country | Investigator | Institution |
|---|---|---|
| Argentina | Chirino Navarta, Daniel | Mautalen Salud e Investigación (Expertia SA), Buenos Aires |
| Argentina | Llapur, Conrado | Clinica Mayo de UMCB SRL, San Miguel de Tucumán |
| Argentina | Maulen, Sergio | Fundación FunDaMos, Buenos Aires |
| Argentina | Perez Marc, Gonzalo | Hospital Militar Central Cirujano Mayor Dr. Cosme Argerich, Buenos Aires |
| Argentina | Polack, Fernando | Hospital Militar Central Cirujano Mayor Dr. Cosme Argerich, Buenos Aires |
| Argentina | Riera, Fernando | Sanatorio Allende, Cordoba |
| Brazil | Diaz, Ricardo | Azidus Brasil Pesquisa e Desenvolvimento Ltda, Valinhos, Sao Paulo |
| Brazil | Ferrara, Luciana | Azidus Brasil Pesquisa e Desenvolvimento Ltda, Valinhos, Sao Paulo |
| Brazil | Freire, Antonio | Santa Casa De Misericordia De Belo Horizonte, Belo Horizonte, Minas Gerais |
| Brazil | Hammes, Luciano | Centro de Pesquisa Clinica - Hospital Moinhos de Vento, Porto Alegre, Rio Grande do Sul |
| Brazil | Itinose, Kengi | Unidade Hospital do Rocio, Campo Largo |
| Brazil | Nogueira, Mauricio | Fundação Faculdade Regional de Medicina de Sao Jose do Rio Preto, Sao Jose do Rio Preto |
| Brazil | Vasconcellos, Eduardo Freire | Instituto de Pesquisas Clinicas L2IP, Brasilia, DF |
| Brazil | Wolff, Priscila Geller | IBPClin Instituto Brasil de Pequisa Clinica, Rio de Janeiro |
| Canada | Abitbol, Alexander | LMC Clinical Research Inc. (CPU), Toronto, Ontario |
| Canada | Aggarwal, Naresh | Aggarwal and Associates Ltd, Brampton, Ontario |
| Canada | Badenhorst, Josias | CARe Clinic, Red Deer, Alberta |
| Canada | Dionne, Marc | CHU de Québec-Université Laval, Quebec City, Quebec |
| Canada | Frechette, Andre | Diex Research Quebec Inc, Quebec City, Quebec |
| Canada | Gauthier, Jean Sebastien | QT Research Sherbrooke, Sherbrooke, Quebec |
| Canada | Girard, Ginette | Diex Research, Sherbrooke, Quebec |
| Canada | Henein, Sam | SKDS Research Inc., Newmarket, Ontario |
| Canada | Libman, Michael | McGill University Health Centre Vaccine Study Centre, Pierrefonds, Quebec |
| Canada | Paquette, Jean-Sebastien | Diex Recherche Joliette, Saint-Charles- |

| Country | Investigator | Institution |
|---|---|---|
| | | Borromée, Quebec |
| Canada | Tellier, Guy | Manna Research Mirabel, Mirabel, Quebec |
| Canada | Vale, Noah | Manna Research Toronto, Toronto, Ontario |
| Canada | Vallieres, Gerald | Manna Research Quebec, Lévis, Quebec |
| Canada | Wallace, Garry | Dawson Clinical Research Inc., Guelph, Ontario |
| Mexico | Castro Melchor, Liliana | Investigación Biomédica para el Desarrollo de Fármacos, S.A. de C.V., Zapopan |
| Mexico | De los Rios Ibarra, Manuel | Centro para el Desarrollo de la Medicina y de Asistencia Médica Especializada SC, Culiacan |
| Mexico | Del Carpio, Luis | Sociedad de Metabolismo y Corazon S.C (SOMECO), Veracruz |
| Mexico | Guzman Romero, Ana Karla | RM Pharma Specialists S.A. de C.V., Ciudad de Mexico |
| Mexico | Martinez Perez, Jose Miguel | Integra RGH Centro de Investigacion - Clinica de Ozonoterapia RGH AC, Puebla |
| Mexico | Rios Mejia, Edmundo Daniel | Investigación Biomédica para el Desarrollo de Fármacos, S.A. de C.V., Aguascalientes |
| Mexico | Sanchez Campos, Efren Alberto | Centro Multidisciplinario para el Desarrollo Especializado de la Investigación Clínica en Yucatan S.C.P. (CEMDEICY S.C.P.), Merida |
| United Kingdom | Apoola, Ade | University Hospitals Derby and Burton, Derby |
| United Kingdom | Galloway, James | Kings College Hospital, London |
| United Kingdom | Healy, Brendan | Public Health Wales NHS Trust, Cardiff |
| United Kingdom | Kyriakidou, Christina | Synexus Midlands Clinical Research Centre, Birmingham |
| United Kingdom | Lacey, Charles | University of York/York Teaching Hospital, York |
| United Kingdom | Moore, Patrick | University Hospitals Dorset NHS Foundation Trust, Bournemouth |
| United Kingdom | Nicholls, Helen | Synexus Wales Clinical Research Centre DRS, Cardiff |
| United Kingdom | Prestwell, Carol | Synexus Lancashire Clinical Research Centre DRS, Chorley |
| United Kingdom | Soiza, Roy | NHS Grampian, Aberdeen |
| United Kingdom | Viljoen, Marlies | Synexus Manchester Clinical Research Centre DRS, Manchester |
| United Kingdom | Whittington, Ashley | London North West University Healthcare NHS Trust, Harrow |
| United States | Aazami, Hessam | Hope Clinical, Canoga Park, California |

| Country | Investigator | Institution |
|---|---|---|
| United States | Adelglass, Jeffrey | Research Your Health, Plano, Texas |
| United States | Anderson, Duane | Excel Clinical Research, Las Vegas, Nevada |
| United States | Arora, Samir | Aventiv Research Inc, Columbus, Ohio |
| United States | Berger, Ira | Meridian Clinical Research, Rockville, Maryland |
| United States | Bordon, Jose | Ascension Providence Health System, Washington, DC |
| United States | Bradley, Paul | Meridian Clinical Research, Savannah, Georgia |
| United States | Carlson, Mark | Be Well Clinical Studies, LLC, Lincoln, Nebraska |
| United States | Casey, Sherri | Benchmark Research, Covington, Louisiana |
| United States | Chu, Laurence | Benchmark Research, Austin, Texas |
| United States | DeFrancisco, Don | Pharmacology Research Institute, Newport Beach, California |
| United States | Ebuh, Valentine | Global Medical Research, Dallas, Texas |
| United States | Ensz, David | Meridian Clinical Research, Sioux City, Iowa |
| United States | Essink, Brandon | Meridian Clinical Research LLC, Omaha, Nebraska |
| United States | Fiel, Thomas | Fiel Family & Sports Medicine/ CCT Research, Tempe, Arizona |
| United States | Forte, Dana | Forte Family Practice/ CCT Research, Las Vegas, Nevada |
| United States | Fussell, Suzanne | Long Beach Clinical Trial Services Inc, Long Beach, California |
| United States | Gregerson, Gary | ASR, LLC, Nampa, Idaho |
| United States | Haffizulla, Jason | Precision Clinical Research, LLC, Sunrise, Florida |
| United States | Hassman, Michael | Hassman Research Institute, Berlin, New Jersey |
| United States | Hemmersmeier, John | South Ogden Family Medicine/ CCT Research, Ogden, Utah |
| United States | Hong, Mathew | M3 Wake Research, Inc, Raleigh, North Carolina |
| United States | Juvvadi, Nikhila | Affinity Health, Chicago, Illinois |
| United States | Latorre, Agustin | AppleMed Research Inc, Miami, Florida |
| United States | Martin, Earl | DM Clinical Research/Martin Diagnostic Clinic, Tomball, Texas |
| United States | Oppong, Yaa | Carolina Institute for Clinical Research, Fayetteville, North Carolina |
| United States | Patel, Suchet | Meridian Clinical Research, LLC, Endwell, New York |
| United States | Pickrell, Paul | Tekton Research, Austin, Texas |
| United States | Rosen, Jeffrey | AMR Miami, formerly Clinical Research of South Florida, Coral Gables, Florida |

| Country | Investigator | Institution |
|---|---|---|
| United States | Rothenberg, Joan | Velocity Clinical Research, Cleveland, Ohio |
| United States | Schwartz, Howard | Research Centers of America, Hollywood, Florida |
| United States | Seger, William | Benchmark Research, Fort Worth, Texas |
| United States | Tarakjian, Denis | WR-MCCR, LLC, San Diego, California |
| United States | Thakkar, Harish | Mt. Olympus Medical Research LLC, Sugarland, Texas |
| United States | Vanderzyl, John | Sugar Lakes Family Practice, PA - an Elligo Health Research Site, Sugarland, Texas |
| United States | Vrbicky, Keith | Meridian Clinical Research LLC, Norfolk, Nebraska |
| United States | Walters, Alton | Las Vegas Clinical Trials, North Las Vegas, Nevada |
| United States | Williams, Barton | Trial Management Associates, LLC, Wilmington, North Carolina |
| United States | Williams, Hayes | Achieve Clinical Research, LLC dba Accel Research Sites at Birmingham Clinical Research Unit, Birmingham, Alabama |
| United States | Wolf, Thomas | Methodist Physicians Clinic / CCT Research, Fremont, Nebraska |
| United States | Zeig, Steven | Elixia LLC, Palm Beach, Florida |

*Supplementary Methods (Viral Sequencing)*

Sequencing of viral genomes from swabs was performed by Viroclinics-DDL (Netherlands). Total nucleic acid (DNA/RNA) isolation was performed with the MagMAX™ Viral/Pathogen Nucleic Acid Isolation kit on the KingFisher™ Flex instrument (ThermoFisher) using 400 µL sample input. Nucleic acid elution was performed in 50 µL of elution buffer. Full-length amplification of the target RNA, i.e., RNA encoding SARS-CoV-2 S, was performed by amplifying 5 overlapping fragments of ~900 bp by nested RT-PCR using the primers sets from the CDC Singleplex PCR 38 amplicons protocol (PacBio). The outer one-step RT-PCR reaction was performed with the outer S-gene primer sets and the SuperScript™ III One-Step RT-PCR System with Platinum™ Taq High Fidelity DNA Polymerase (ThermoFisher) using 5 µL of the isolated RNA. The nested (inner) amplification step was performed with the inner primer sets and the Phusion Hot Start II High-Fidelity DNA Polymerase (ThermoFisher) or Q5® Hot Start High-Fidelity DNA Polymerase (NEB). Nested PCR products were analyzed by gel electrophoresis using standard molecular biology protocols, to confirm successful amplification.

Next-generation sequence (NGS) analysis of S-gene amplicons was performed using the MiSeq or NextSeq Illumina platforms. NGS analysis was performed on samples consisting of two pools of non-overlapping S-gene amplicons. To generate single datasets for reporting and interpretation, the FASTQ files of the two pools of non-overlapping fragments were pooled together *in silico*. After combining the two datasets, all data was mapped against the same reference sequence (SARS-CoV-2 Wuhan-Hu-1 GenBank: MN908947.3). Identifying the location on the reference genome allowed identification of reads that started or ended in or near a primer region (possibly containing primer ambiguity), in order to select them for the primer trimming algorithm. Sufficient nucleotides were trimmed off each read that was selected by this process and the remaining part of the read was considered non-primer derived. For *in silico* pooling and primer trimming, all primer locations were implemented in DDL's bioinformatics ATHENA pipeline. The deep sequencing process comprised: quantification of S-gene amplicon pools by PicoGreen or Qubit™; fragment dilutions and equimolar mixing to normalize the concentrations of each of the amplicon pools; library preparation with the Nextera XT System (Illumina); sequence reactions with 300-cycle kit (150 bp bidirectional reads) on the Illumina Miseq or with a 300-cycle mid output kit on the Illumina Nextseq; Data QC and Data reporting.

*Supplementary Figure 1 | Circulating Variants During Course of Clinical Trial*

Each panel illustrates the variants circulating in time relative to the accrual of COVID-19 events for each nation in which the trial took place. Individual vertical columns indicate the incidence rate of variants reported to GISAID during a seven-day period relative to the total number of sequences reported for the period. Individual colors within the stacked columns represent variants in circulation as a percentage of total, shown on the left Y axis. In cases where sequence identity was not reported or does not refer to a Variant of Concern (VoC) or Variant of Interest (VoI), the incidence was reported as undefined. Red lines indicate the number of total adjudicated COVID-19 events contributing to the calculation of primary vaccine efficacy accrued in each nation through time quantified relative to the right Y-axis. No COVID-19 events accrued in Mexico. Dotted vertical lines indicate the date of first injection of a participant by country. FPI: First Patient In.

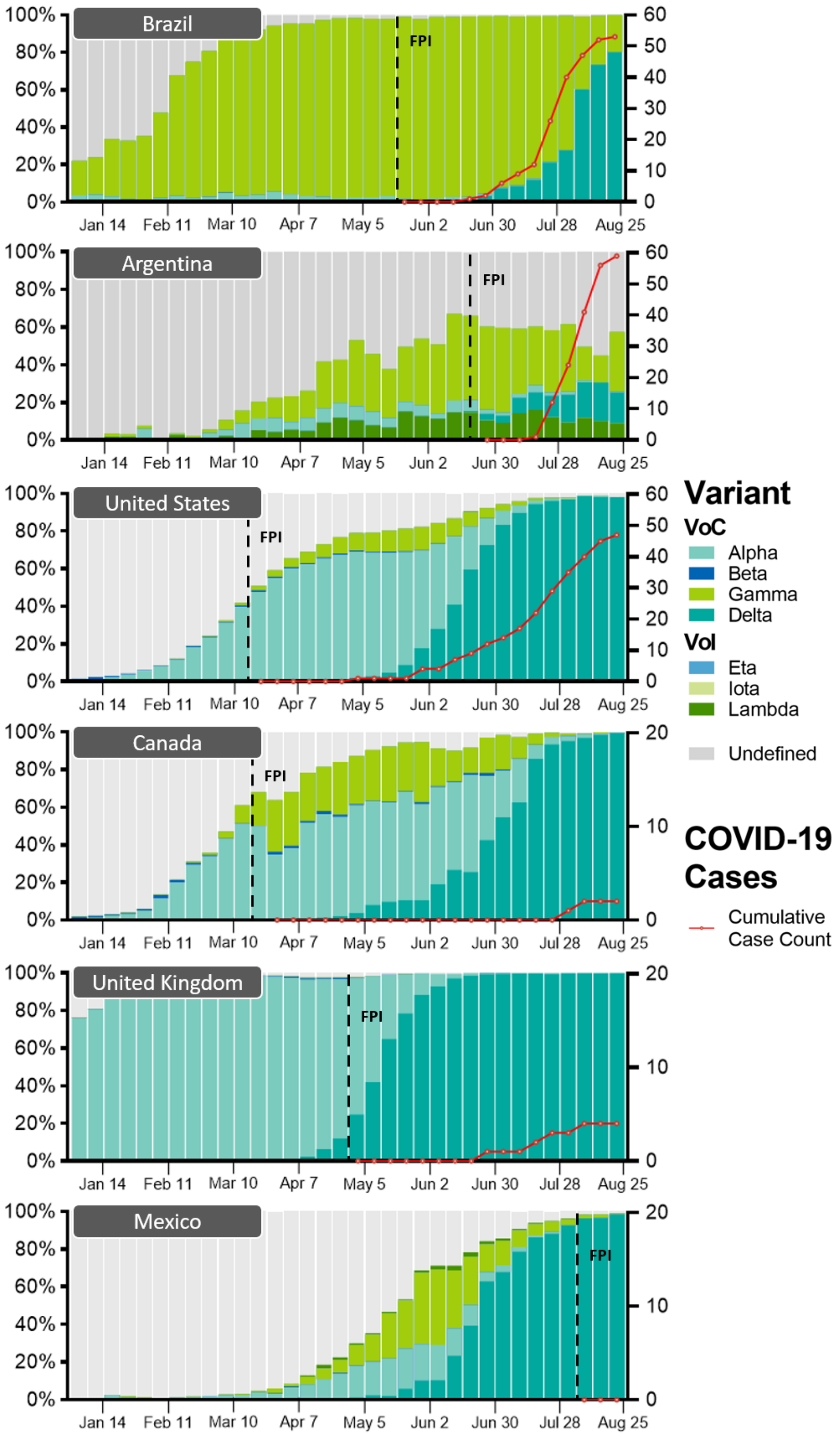
Brazil
Argentina
United States
Canada
United Kingdom
Mexico
FPI
100%
80%
60%
40%
20%
0%
Jan 14
Feb 11
Mar 10
Apr 7
May 5
Jun 2
Jun 30
Jul 28
Aug 25
Variant
VoC
Alpha
Beta
Gamma
Delta
VoI
Eta
Iota
Lambda
Undefined
COVID-19 Cases
Cumulative Case Count

*Supplementary Table S1 | Summary of demographics in the ITT set (NCT04636697)*

'N' is the number of participants in a population, while 'n' represents the number of participants in a category. Race or ethnic group was reported by the participants (who could have listed more than one category). SD: Standard deviation; Min.: Minimum; Max.: Maximum

| | **Healthy Adult** | | **Older Adults (65+)** | | **Adults with Comorbidities** | | **All Participants** | |
|---|---|---|---|---|---|---|---|---|
| | **CoVLP + AS03** | **Placebo** | **CoVLP + AS03** | **Placebo** | **CoVLP + AS03** | **Placebo** | **CoVLP + AS03** | **Placebo** |
| **Participants (N)** | 10,816 | 10,835 | 62 | 65 | 1195 | 1166 | 12,074 | 12,066 |
| **Sex, n (%)** | | | | | | | | |
| Male | 5,460 (50.5) | 5,553 (51.3) | 35 (56.5) | 35 (53.8) | 612 (51.2) | 598 (51.3) | 6,107 (50.6) | 6,186 (51.3) |
| Female | 5,356 (49.5) | 5,282 (48.7) | 27 (43.5) | 30 (46.2) | 583 (48.8) | 568 (48.7) | 5,966 (49.4) | 5,880 (48.7) |
| **Race, n (%)** | | | | | | | | |
| American Indian/ Alaska Native | 27 (0.2) | 23 (0.2) | 0 | 0 | 3 (0.3) | 7 (0.6) | 30 (0.2) | 30 (0.2) |
| Asian | 137 (1.3) | 137 (1.3) | 1 (1.6) | 0 | 11 (0.9) | 10 (0.9) | 149 (1.2) | 147 (1.2) |
| Black/ African American | 681 (6.3) | 667 (6.2) | 8 (12.9) | 13 (20.0) | 160 (13.4) | 155 (13.3) | 849 (7.0) | 835 (6.9) |
| Multiple | 197 (1.8) | 237 (2.2) | 1 (1.6) | 0 | 26 (2.2) | 26 (2.2) | 224 (1.9) | 263 (2.2) |
| Native Hawaiian or Other Pacific Islander | 24 (0.2) | 21 (0.2) | 0 | 0 | 2 (0.2) | 4 (0.3) | 26 (0.2) | 25 (0.2) |
| White or Caucasian | 9,694 (89.6) | 9,690 (89.4) | 51 (82.3) | 51 (78.5) | 981 (82.1) | 959 (82.2) | 10,726 (88.8) | 10,700 (88.7) |
| Other | 1 (<0.1) | 2 (<0.1) | 0 | 0 | 0 | 0 | 1 (<0.1) | 2 (<0.1) |
| Not Reported | 52 (0.5) | 54 (0.5) | 0 | 1 (1.5) | 10 (0.8) | 5 (0.4) | 62 (0.5) | 60 (0.5) |
| **Ethnicity, n (%)** | | | | | | | | |
| Hispanic or Latinx | 8,965 (82.9) | 8,955 (82.6) | 15 (24.2) | 26 (40.0) | 915 (76.6) | 915 (78.5) | 9,895 (82.0) | 9,896 (82.0) |
| Not Hispanic or Latinx | 1,800 (16.6) | 1,830 (16.9) | 47 (75.8) | 39 (60.0) | 273 (22.8) | 241 (20.7) | 2,120 (17.6) | 2,110 (17.5) |
| Not Reported/ Specified | 51 (0.5) | 50 (0.5) | 0 | 0 | 7 (0.6) | 10 (0.9) | 58 (0.5) | 60 (0.5) |
| **Age at consent (years)** | | | | | | | | |
| Mean (SD) | 32.0 (11.07) | 31.9 (11.02) | 69.9 (4.15) | 69.5 (5.00) | 38.8 (13.72) | 39.2 (14.14) | 32.8 (11.82) | 32.8 (11.85) |
| Median | 29.0 | 29.0 | 69.0 | 68.0 | 37.0 | 38.0 | 29.0 | 29.0 |
| Min, Max | 18, 64 | 18, 64 | 65, 80 | 65, 86 | 18, 82 | 18, 86 | 18, 82 | 18, 86 |

*Supplementary Table S2 | Baseline Comorbidities*

'N' is the total number of participants who received a given vaccination. 'n' is the number of a participants who reported the specified comorbid condition and is shown as a percentage in brackets. Comorbidities reported for ≤0.1% of the total population have not been included.

| **n (%)** | **CoVLP+AS03 N = 12036** | **Placebo N = 12040** | **Total N = 24076** |
|---|---|---|---|
| **Participants with at least one comorbidity** | **1781 (14.8)** | **1686 (14.0)** | **3467 (14.4)** |
| Obesity | 1246 (10.4) | 1171 (9.7) | 2417 (10.0) |
| Documented Hypertension | 340 (2.8) | 322 (2.8) | 672 (2.8) |
| Type 2 Diabetes | 116 (1.0) | 100 (0.8) | 216 (0.9) |
| Chronic Obstructive Lung disease (COPD) | 41 (0.3) | 45 (0.4) | 86 (0.4) |
| Immunocompromised | 48 (0.4) | 40 (0.3) | 88 (0.4) |
| Cardiovascular disease | 40 (0.3) | 41 (0.3) | 81 (0.3) |
| Asthma | 25 (0.2) | 17 (0.1) | 42 (0.2) |

*Supplementary Table S3 | Efficacy by Subgroup (ITT)*

For most subgroups (1), values indicate the number of positive cases (total number of participants in the subgroup: percentage positivity). For variants (2), values indicate number of sequenced cases assigned to the variant (percent positive). Yrs indicate years, 95% CI indicates 95% confidence interval. VE indicates Vaccine Efficacy and NA indicates not applicable.

| Subgroup | Placebo | CoVLP+AS03 | VE (95% CI) |
|---|---|---|---|
| **All Participants** | 125 (12,067; 1.0%) | 40 (12,074; 0.3%) | 69.5 (56.7, 78.8) |
| **Baseline Health, Age** | | | |
| Healthy, ≥18 to <65 yrs | 111 (10,835, 1.0%) | 36 (10,0816, 0.3%) | 68.9 (55.0, 78.9) |
| Healthy, ≥65 yrs | 1 (65; 1.5%) | 1 (62, 1.6%) | 12.9 (-3,295.5, 97.8) |
| Significant comorbidities, ≥18 yrs | 13 (1,166; 1.1%) | 3 (1,195, 0.3%) | 78.6 (30.2, 95.1) |
| **Sex** | | | |
| Male | 58 (6,186; 0.9%) | 22 (6,107, 0.4%) | 63.5 (41.0, 78.1) |
| Female | 67 (5,880; 1.1%) | 18 (5,966, 0.3%) | 74.5 (57.8, 85.2) |
| **Race or Ethnic Group** | | | |
| Asian | 2 (147; 1.4%) | 0 (149; 0%) | 100.0 (-195.1, NA) |
| Black or African American | 9 (835; 1.1%) | 5 (849; 0.6%) | 47.3 (-57.6, 84.0) |
| White or Caucasian | 107 (10,700; 1.0%) | 34 (10,726; 0.3%) | 69.7 (55.7, 79.6) |
| Latinx or Hispanic | 88 (9,896; 0.9%) | 31 (9,895; 0.3%) | 66.6 (50.1, 78.1) |
| Not Latinx or Hispanic | 37 (2,110; 1.8%) | 9 (2,120; 0.4%) | 76.6 (52.8, 89.3) |
| **Disease Severity** | | | |
| Moderate | 33 (12,067; 0.3%) | 8 (12,074; <0.1%) | 76.8 (51.4, 90.0) |
| Moderate/Severe | 36 (12,067; 0.3%) | 8 (12,074; <0.1%) | 78.8 (55.8, 90.8) |
| Severe | 3 (12,067; <0.1%) | 0 (12,074; 0%) | 100.0 (-63.7, NA) |
| **Seropositivity at Baseline** | | | |
| Seropositive | 3 (1,791; 0.2%) | 6 (1,776; 0.3%) | -94.3 (-852.1, 51.6) |
| Seronegative | 121 (10,106; 1.2%) | 33 (10,115; 0.3%) | 74.0 (62.1, 82.5) |
| **Variants** | | | |
| Ancestral | 0 (0%) | 0 (0%) | NA |
| Alpha | 6 (<0.1%) | 0 (0%) | 100 (38.2 NA) |
| Beta | 0 (0%) | 0 (0%) | NA |
| Gamma | 47 (0.4%) | 6 (<0.1%) | 87.8 (73.0, 95.3) |
| Delta | 44 (0.4%) | 12 (<0.1%) | 74.0 (51.7, 86.8) |
| Lambda | 3 (<0.1%) | 0 (0%) | 100 (-63.7, NA) |
| Mu | 4 (<0.1%) | 0 (0%) | 100.0 (-6.4, NA) |
| Omicron | 0 (0%) | 0 (0%) | NA |

*Supplementary Table S4 | Incidence of Solicited Local Adverse Events (Safety Analysis Set)*

'N' is the total number of participants in the Safety Analysis Set who received a given vaccination. 'n' is the number of a participants reporting at least 1 occurrence of the specified event category and is shown as a percentage in brackets. For each category, Grade 1 = Mild; Grade 2 = Moderate; Grade 3 = Severe; Grade 4 = Potentially life threatening. If a participant had the same AE but with different grades over time, the highest grade was reported. 'Missing' rows counted participants within the population if they did not have a non-missing assessment or had an assessment with a non-graded value.

| n (%) | Vaccination 1 | | Vaccination 2 | | Overall | |
|---|---|---|---|---|---|---|
| | CoVLP+ AS03<br>N = 4 136 | Placebo<br>N = 3 683 | CoVLP+ AS03<br>N = 4 136 | Placebo<br>N = 3 683 | CoVLP+ AS03<br>N = 4 136 | Placebo<br>N = 3 683 |
| **At least 1 solicited local AE** | **3538 (85.5)** | **1149 (31.2)** | **3496 (84.5)** | **1135 (30.8)** | **3819 (92.3)** | **1677 (45.5)** |
| Grade 1 | 3021 (73.0) | 1123 (30.5) | 2494 (60.3) | 1111 (30.2) | 2590 (62.6) | 1629 (44.2) |
| Grade 2 | 484 (11.7) | 24 (0.7) | 917 (22.2) | 23 (0.6) | 1115 (27.0) | 45 (1.2) |
| Grade 3 | 33 (0.8) | 2 (<0.1) | 85 (2.1) | 1 (<0.1) | 114 (2.8) | 3 (<0.1) |
| Grade 4 | 0 | 0 | 0 | 0 | 0 | 0 |
| Missing | 4 (<0.1) | 6 (0.2) | 74 (1.8) | 12 (0.3) | 2 (<0.1) | 5 (0.1) |
| **Pain** | **3514 (85.0)** | **1 082 (29.4)** | **3 483 (84.2)** | **1 100 (29.9)** | **3 806 (92.0)** | **1 604 (43.6)** |
| Grade 1 | 3 072 (74.3) | 1 066 (28.9) | 2 625 (63.5) | 1 080 (29.3) | 2 758 (66.7) | 1 570 (42.6) |
| Grade 2 | 420 (10.2) | 15 (0.4) | 830 (20.1) | 20 (0.5) | 999 (24.2) | 33 (0.9) |
| Grade 3 | 22 (0.5) | 1 (<0.1) | 28 (0.7) | 0 | 49 (1.2) | 1 (<0.1) |
| Grade 4 | 0 | 0 | 0 | 0 | 0 | 0 |
| Missing | 4 (<0.1) | 6 (0.2) | 74 (1.8) | 12 (0.3) | 2 (<0.1) | 5 (0.1) |
| **Redness** | **281 (6.8)** | **107 (2.9)** | **633 (15.3)** | **77 (2.1)** | **815 (19.7)** | **167 (4.5)** |
| Grade 1 | 238 (5.8) | 101 (2.7) | 446 (10.8) | 75 (2.0) | 596 (14.4) | 159 (4.3) |
| Grade 2 | 36 (0.9) | 5 (0.1) | 145 (3.5) | 1 (<0.1) | 172 (4.2) | 6 (0.2) |
| Grade 3 | 7 (0.2) | 1 (<0.1) | 42 (1.0) | 1 (<0.1) | 47 (1.1) | 2 (<0.1) |
| Grade 4 | 0 | 0 | 0 | 0 | 0 | 0 |
| Missing | 6 (0.1) | 8 (0.2) | 76 (1.8) | 13 (0.4) | 3 (<0.1) | 5 (0.1) |
| **Swelling** | **774 (18.7)** | **108 (2.9)** | **1241 (30.0)** | **113 (3.1)** | **1548 (37.4)** | **200 (5.4)** |
| Grade 1 | 660 (16.0) | 100 (2.7) | 974 (23.5) | 110 (3.0) | 1211 (29.3) | 189 (5.1) |
| Grade 2 | 102 (2.5) | 7 (0.2) | 240 (5.8) | 3 (<0.1) | 300 (7.3) | 10 (0.3) |
| Grade 3 | 12 (0.3) | 1 (<0.1) | 27 (0.7) | 0 | 37 (0.9) | 1 (<0.1) |
| Grade 4 | 0 | 0 | 0 | 0 | 0 | 0 |
| Missing | 5 (0.1) | 6 (0.2) | 74 (1.8) | 12 (0.3) | 2 (<0.1) | 5 (0.1) |

*Supplementary Table S5 | Incidence of Solicited Systemic Adverse Events (Safety Analysis Set)*

'N' is the total number of participants in the Safety Analysis Set who received a given vaccination. 'n' is the number of a participants reporting at least 1 occurrence of the specified event category and is shown as a percentage in brackets. For each category, Grade 1 = Mild; Grade 2 = Moderate; Grade 3 = Severe; Grade 4 = Potentially life threatening. If a participant had the same AE but with different grades over time, the highest grade was reported. 'Missing' rows counted participants within the population if they did not have a non-missing assessment or had an assessment with a non-graded value. Medically implausible fever temperature values were excluded from table.

| **n (%)** | **Vaccination 1** | | **Vaccination 2** | | **Overall** | |
|---|---|---|---|---|---|---|
| | **CoVLP+ AS03** | **Placebo** | **CoVLP+ AS03** | **Placebo** | **CoVLP+ AS03** | **Placebo** |
| | N = 4 136 | N = 3 683 | N = 4 136 | N = 3 683 | N = 4 136 | N = 3 683 |
| **At least 1 solicited systemic AE** | **2835 (68.5)** | **2023 (54.9)** | **3190 (77.1)** | **1517 (41.2)** | **3612 (87.3)** | **2394 (65.0)** |
| Grade 1 | 2017 (48.8) | 1602 (43.5) | 1388 (33.6) | 1147 (31.1) | 1538 (37.2) | 1734 (47.1) |
| Grade 2 | 776 (18.8) | 397 (10.8) | 1671 (40.4) | 349 (9.5) | 1906 (46.1) | 617 (16.8) |
| Grade 3 | 42 (1.0) | 24 (0.7) | 129 (3.1) | 20 (0.5) | 166 (4.0) | 42 (1.1) |
| Grade 4 | 0 | 0 | 2 (<0.1) | 1 (<0.1) | 2 (<0.1) | 1 (<0.1) |
| Missing | 2 (<0.1) | 6 (0.2) | 72 (1.7) | 12 (0.3) | 2 (<0.1) | 5 (0.1) |
| **Chills** | **571 (13.8)** | **352 (9.6)** | **1 693 (40.9)** | **263 (7.1)** | **1 889 (45.7)** | **522 (14.2)** |
| Grade 1 | 469 (11.3) | 314 (8.5) | 966 (23.4) | 228 (6.2) | 1 108 (26.8) | 451 (12.2) |
| Grade 2 | 98 (2.4) | 38 (1.0) | 702 (17.0) | 34 (0.9) | 753 (18.2) | 70 (1.9) |
| Grade 3 | 4 (<0.1) | 0 | 24 (0.6) | 1 (<0.1) | 27 (0.7) | 1 (<0.1) |
| Grade 4 | 0 | 0 | 1 (<0.1) | 0 | 1 (<0.1) | 0 |
| Missing | 8 (0.2) | 9 (0.2) | 74 (1.8) | 12 (0.3) | 2 (<0.1) | 5 (0.1) |
| **Fatigue** | **1 490 (36.0)** | **1 038 (28.2)** | **2 277 (55.1)** | **792 (21.5)** | **2 634 (63.7)** | **1 352 (36.7)** |
| Grade 1 | 1 144 (27.7) | 820 (22.3) | 1 270 (30.7) | 638 (17.3) | 1 460 (35.3) | 1 018 (27.6) |
| Grade 2 | 332 (8.0) | 206 (5.6) | 942 (22.8) | 142 (3.9) | 1 096 (26.5) | 311 (8.4) |
| Grade 3 | 14 (0.3) | 12 (0.3) | 65 (1.6) | 12 (0.3) | 78 (1.9) | 23 (0.6) |
| Grade 4 | 0 | 0 | 0 | 0 | 0 | 0 |
| Missing | 8 (0.2) | 8 (0.2) | 72 (1.7) | 12 (0.3) | 2 (<0.1) | 5 (0.1) |
| **Feeling of General Discomfort** | **1 200 (29.0)** | **699 (19.0)** | **2280 (55.1)** | **574 (15.6)** | **2 595 (62.7)** | **1002 (27.2)** |
| Grade 1 | 907 (21.9) | 582 (15.8) | 1 154 | 459 (12.5) | 1 341 | 784 (21.3) |

| n (%) | Vaccination 1 | | Vaccination 2 | | Overall | |
|---|---|---|---|---|---|---|
| | CoVLP+ AS03 | Placebo | CoVLP+ AS03 | Placebo | CoVLP+ AS03 | Placebo |
| | N = 4 136 | N = 3 683 | N = 4 136 | N = 3 683 | N = 4 136 | N = 3 683 |
| | | | (27.9) | | (32.4) | |
| Grade 2 | 278 (6.7) | 111 (3.0) | 1 089 (26.3) | 109 (3.0) | 1 203(29.1 ) | 207 (5.6) |
| Grade 3 | 15 (0.4) | 6 (0.2) | 36 (0.9) | 6 (0.2) | 50 (1.2) | 11 (0.3) |
| Grade 4 | 0 | 0 | 1 (<0.1) | 0 | 1 (<0.1) | 0 |
| Missing | 8 (0.2) | 9 (0.2) | 74 (1.8) | 13 (0.4) | 2 (<0.1) | 5 (0.1) |
| **Feeling of Swelling in the Axilla (Armpit)** | **266 (6.4)** | **123 (3.3)** | **461 (11.1)** | **71 (1.9)** | **613 (14.8)** | **182 (4.9)** |
| Grade 1 | 233 (5.6) | 121 (3.3) | 389 (9.4) | 66 (1.8) | 515 (12.5) | 175 (4.8) |
| Grade 2 | 32 (0.8) | 2 (<0.1) | 70 (1.7) | 5 (0.1) | 95 (2.3) | 7 (0.2) |
| Grade 3 | 1 (<0.1) | 0 | 2 (<0.1) | 0 | 3 (<0.1) | 0 |
| Grade 4 | 0 | 0 | 0 | 0 | 0 | 0 |
| Missing | 8 (0.2) | 9 (0.2) | 75 (1.8) | 13 (0.4) | 3 (<0.1) | 5 (0.1) |
| **Feeling of Swelling in the Neck** | **475 (11.5)** | **328 (8.9)** | **632 (15.3)** | **245 (6.7)** | **905 (21.9)** | **484 (13.1)** |
| Grade 1 | 427 (10.3) | 298 (8.1) | 514 (12.4) | 217 (5.9) | 746 (18.0) | 432 (11.7) |
| Grade 2 | 46 (1.1) | 28 (0.8) | 112 (2.7) | 28 (0.8) | 151 (3.7) | 50 (1.4) |
| Grade 3 | 2 (<0.1) | 2 (<0.1) | 6 (0.1) | 0 | 8 (0.2) | 2 (<0.1) |
| Grade 4 | 0 | 0 | 0 | 0 | 0 | 0 |
| Missing | 8 (0.2) | 9 (0.2) | 75 (1.8) | 13 (0.4) | 3 (<0.1) | 5 (0.1) |
| **Fever** | **44 (1.1)** | **34 (0.9)** | **355 (8.6)** | **18 (0.5)** | **390 (9.4)** | **52 (1.4)** |
| Grade 1 | 31 (0.7) | 26 (0.7) | 259 (6.3) | 10 (0.3) | 282 (6.8) | 36 (1.0) |
| Grade 2 | 10 (0.2) | 7 (0.2) | 70 (1.7) | 6 (0.2) | 79 (1.9) | 13 (0.4) |
| Grade 3 | 3 (<0.1) | 1 (<0.1) | 25 (0.6) | 2 (<0.1) | 28 (0.7) | 3 (<0.1) |
| Grade 4 | 0 | 0 | 1 (<0.1) | 0 | 1 (<0.1) | 0 |
| Missing | 59 (1.4) | 59 (1.6) | 92 (2.2) | 27 (0.7) | 8 (0.2) | 11 (0.3) |
| **Headache** | **1 644 (39.7)** | **1 299 (35.3)** | **2 325 (56.2)** | **940 (25.5)** | **2 786 (67.4)** | **1 662 (45.1)** |
| Grade 1 | 1 288 (31.1) | 1 089 (29.6) | 1 363 (33.0) | 732 (19.9) | 1 634 (39.5) | 1300 (35.3) |
| Grade 2 | 342 (8.3) | 200 (5.4) | 927 (22.4) | 196 (5.3) | 1 105 (26.7) | 341 (9.3) |
| Grade 3 | 14 (0.3) | 10 (0.3) | 34 (0.8) | 11 (0.3) | 46 (1.1) | 20 (0.5) |
| Grade 4 | 0 | 0 | 1 (<0.1) | 1 (<0.1) | 1 (<0.1) | 1 (<0.1) |
| Missing | 6 (0.1) | 9 (0.2) | 74 (1.8) | 13 (0.4) | 2 (<0.1) | 5 (0.1) |
| **Joint Aches** | **642 (15.5)** | **406 (11.0)** | **1 361 (32.9)** | **313 (8.5)** | **1 627 (39.3)** | **595 (16.2)** |
| Grade 1 | 510 (12.3) | 355 (9.6) | 838 (20.3) | 268 (7.3) | 1 028 (24.9) | 502 (13.6) |
| Grade 2 | 129 (3.1) | 47 (1.3) | 504 (12.2) | 44 (1.2) | 578 (14.0) | 88 (2.4) |
| Grade 3 | 3 (<0.1) | 4 (0.1) | 19 (0.5) | 1 (<0.1) | 21 (0.5) | 5 (0.1) |
| Grade 4 | 0 | 0 | 0 | 0 | 0 | 0 |

| n (%) | Vaccination 1 | | Vaccination 2 | | Overall | |
|---|---|---|---|---|---|---|
| | CoVLP+ AS03 | Placebo | CoVLP+ AS03 | Placebo | CoVLP+ AS03 | Placebo |
| | N = 4 136 | N = 3 683 | N = 4 136 | N = 3 683 | N = 4 136 | N = 3 683 |
| Missing | 8 (0.2) | 8 (0.2) | 72 (1.7) | 12 (0.3) | 2 (<0.1) | 5 (0.1) |
| **Muscle Aches** | **1 768 (42.7)** | **771 (20.9)** | **2 273 (55.0)** | **617 (16.8)** | **2770 (67.0)** | **1 083 (29.4)** |
| Grade 1 | 1 437 (34.7) | 698 (19.0) | 1 354 (32.7) | 539 (14.6) | 1 683 (40.7) | 944 (25.6) |
| Grade 2 | 318 (7.7) | 71 (1.9) | 875 (21.2) | 73 (2.0) | 1 031 (24.9) | 133 (3.6) |
| Grade 3 | 13 (0.3) | 2 (<0.1) | 43 (1.0) | 5 (0.1) | 55 (1.3) | 6 (0.2) |
| Grade 4 | 0 | 0 | 1 (<0.1) | 0 | 1 (<0.1) | 0 |
| Missing | 7 (0.2) | 8 (0.2) | 72 (1.7) | 12 (0.3) | 2 (<0.1) | 5 (0.1) |

*Supplementary Table S6 | Overall Summary of Unsolicited Adverse Events Twenty-One Days after Each Vaccine (Safety Analysis Set)*

'N' is the total number of participants in the Safety Analysis Set who received a given vaccination. 'n' is the number of a participants reporting at least 1 occurrence of the specified event category and is shown as a percentage in brackets. Grade 3 = Severe. Abbreviations: AE, adverse event; AESI, adverse event of special interest; CoVLP, coronavirus-like particle; MAAE, medically attended adverse event; n, number of subjects in category; pIMDs, potential immune-mediated diseases, SAE, serious adverse event; VAED, vaccine-associated enhanced disease.

| **n (%)** | **Vaccination 1** | | **Vaccination 2** | | **Overall** | |
|---|---|---|---|---|---|---|
| | **CoVLP+ AS03** | **Placebo** | **CoVLP+ AS03** | **Placebo** | **CoVLP+ AS03** | **Placebo** |
| | N= 12 036 | N= 12 040 | N= 11 312 | N= 10 775 | N= 12 036 | N= 12040 |
| **Immediate Unsolicited AEs** | 19 (0.2) | 19 (0.2) | 4 (<0.1) | 7 (<0.1) | 23 (0.2) | 26 (0.2) |
| **Unsolicited AEs** | 1 722 (14.3) | 1 647 (13.7) | 1 424 (12.6) | 1 132 (10.5) | 2 735 (22.7) | 2 451 (20.4) |
| ≥ Grade 3 | 39 (0.3) | 38 (0.3) | 30 (0.3) | 30 (0.3) | 69 (0.6) | 65 (0.5) |
| **SAEs** | 13 (0.1) | 9 (<0.1) | 11 (<0.1) | 8 (<0.1) | 24 (0.2) | 16 (0.1) |
| **Related SAEs** | 0 | 0 | 0 | 0 | 0 | 0 |
| **MAAEs** | 287 (2.4) | 276 (2.3) | 242 (2.1) | 230 (2.1) | 509 (4.2) | 490 (4.1) |
| **Related MAAEs** | 28 (0.2) | 21 (0.2) | 20 (0.2) | 22 (0.2) | 48 (0.4) | 43 (0.4) |
| **AEs Leading to Study Withdrawal** | 1 (<0.1) | 5 (<0.1) | 0 | 2 (<0.1) | 1 (<0.1) | 7 (<0.1) |
| **AESIs Reported by the Investigator** | 2 (<0.1) | 0 | 2 (<0.1) | 0 | 4 (<0.1) | 0 |
| **AESIs Identified by Data analysis** | | | | | | |
| Potential VAED | 0 | 0 | 0 | 0 | 0 | 0 |
| Related Potential VAED | 0 | 0 | 0 | 0 | 0 | 0 |
| Related Anaphylactic Reactions | 0 | 0 | 0 | 0 | 0 | 0 |
| pIMDs | 8 (<0.1) | 5 (<0.1) | 7 (<0.1) | 3 (<0.1) | 15 (0.1) | 8 (<0.1) |
| Related pIMDs | 1 (<0.1) | 0 | 1 (<0.1) | 1 (<0.1) | 2 (<0.1) | 1 (<0.1) |
| **AEs Leading to Death** | 0 | 2 (<0.1) | 0 | 2 (<0.1) | 0 | 4 (<0.1) |

*Supplementary Table S7 | Overall Summary of Unsolicited Adverse Events 43 to 201 Days after Second Vaccination (Safety Analysis Set)*

'N' is the total number of participants in the Safety Analysis Set who received a given vaccination. 'n' is the number of a participants reporting at least 1 occurrence of the specified event category and is shown as a percentage in brackets. Grade 3 = Severe. Abbreviations: AE, adverse event; AESI, adverse event of special interest; CoVLP, coronavirus-like particle; MAAE, medically attended adverse event; n, number of subjects in category; pIMDs, potential immune-mediated diseases, SAE, serious adverse event; VAED, vaccine-associated enhanced disease.

| **n (%)** | **Vaccination 1** | | **Vaccination 2** | | **Overall** | |
|---|---|---|---|---|---|---|
| | **CoVLP+ AS03** | **Placebo** | **CoVLP+ AS03** | **Placebo** | **CoVLP+ AS03** | **Placebo** |
| | N= 12 036 | N= 12 040 | N= 11 312 | N= 10 775 | N= 12 036 | N= 12040 |
| **Unsolicited AEs** | 1 (<0.1) | 5 (<0.1) | 510 (4.5) | 475 (4.4) | 511 (4.2) | 480 (4.0) |
| ≥ Grade 3 | 0 | 1 (<0.1) | 21 (0.2) | 28 (0.3) | 21 (0.2) | 29 (0.2) |
| **SAEs** | 0 | 2 (<0.1) | 19 (0.2) | 20 (0.2) | 19 (0.2) | 22 (0.2) |
| **Related SAEs** | 0 | 0 | 0 | 1 (<0.1) | 0 | 1 (<0.1) |
| **MAAEs** | 1 (<0.1) | 3 (<0.1) | 199 (1.8) | 197 (1.8) | 200 (1.7) | 200 (1.7) |
| **Related MAAEs** | 0 | 0 | 2 (<0.1) | 2 (<0.1) | 2 (<0.1) | 2 (<0.1) |
| **AEs Leading to Study Withdrawal** | 1 (<0.1) | 0 | 3 (<0.1) | 5 (<0.1) | 4 (<0.1) | 5 (<0.1) |
| **AESIs Reported by the Investigator** | 0 | 0 | 2 (<0.1) | 0 | 2 (<0.1) | 0 |
| **AESIs Identified by Data analysis** | | | | | | |
| Potential VAED | 0 | 0 | 0 | 0 | 0 | 0 |
| Related Potential VAED | 0 | 0 | 0 | 0 | 0 | 0 |
| Related Anaphylactic Reactions | 0 | 0 | 0 | 0 | 0 | 0 |
| pIMDs | 0 | 0 | 6 (<0.1) | 2 (<0.1) | 6 (<0.1) | 2 (<0.1) |
| Related pIMDs | 0 | 0 | 0 | 0 | 0 | 0 |
| **AEs Leading to Death** | 0 | 0 | 4 (<0.1) | 5 (<0.1) | 4 (<0.1) | 5 (<0.1) |

*Supplementary Table S8 | Incidence of Unsolicited Adverse Events (Safety Analysis Set)*

'N' is the total number of participants in the Safety Analysis Set who received a given vaccination. 'n' is the number of a participants reporting at least 1 occurrence of the specified event PT category and is shown as a percentage in brackets. All events PT reported in ≥1% of subjects after receiving first and or second treatment in CoVLP+AS03 treatment group are listed. PT, preferred term.

| n (%) | Vaccination 1 | | Vaccination 2 | | Overall | |
|---|---|---|---|---|---|---|
| | CoVLP+ AS03 N= 12 036 | Placebo N= 12 040 | CoVLP+ AS03 N= 11 312 | Placebo N= 10 775 | CoVLP+ AS03 N= 12 036 | Placebo N= 12040 |
| **Subjects with at least 1 unsolicited AE** | **1 722 (14.3)** | **1 647 (13.7)** | **1 424 (12.6)** | **1 132 (10.5)** | **2 735 (22.7)** | **2 451 (20.4)** |
| Influenza | 43 (0.4) | 61 (0.5) | 110 (1.0) | 88 (0.8) | 152 (1.3) | 149 (1.2) |
| Headache | 268 (2.2) | 268 (2.2) | 165 (1.5) | 165 (1.5) | 410 (3.4) | 405 (3.4) |
| Nasal congestion | 115 (1.0) | 131 (1.1) | 111 (1.0) | 110 (1.0) | 215 (1.8) | 223 (1.9) |
| Oropharyngeal pain | 76 (0.6) | 90 (0.7) | 75 (0.7) | 82 (0.8) | 147 (1.2) | 169 (1.4) |
| Cough | 57 (0.5) | 73 (0.6) | 60 (0.5) | 57 (0.5) | 115 (1.0) | 127 (1.1) |
| Diarrhea | 61 (0.5) | 61 (0.5) | 72 (0.6) | 54 (0.5) | 129 (1.1) | 110 (0.9) |
| Nausea | 61 (0.5) | 45 (0.4) | 74 (0.7) | 42 (0.4) | 128 (1.1) | 85 (0.7) |
| Myalgia | 90 (0.7) | 67 (0.6) | 62 (0.5) | 43 (0.4) | 146 (1.2) | 109 (0.9) |

*Supplementary Table S9 | Incidence of Adverse Events of Special Interest based on Serious Adverse Events Reported after Vaccination with Other COVID-19 Vaccines (Safety Analysis Set)*

'N' is the total number of participants in the Safety Analysis Set who received a given vaccination. 'n' is the number of a participants reporting at least 1 occurrence of the specified event category and is shown as a percentage in brackets.

| n (%) | Vaccination 1 | | Vaccination 2 | | Overall | |
|---|---|---|---|---|---|---|
| | **CoVLP+ AS03** | **Placebo** | **CoVLP+ AS03** | **Placebo** | **CoVLP+ AS03** | **Placebo** |
| | N= 12 036 | N= 12 040 | N= 11 312 | N= 10 775 | N= 12 036 | N= 12040 |
| Acute respiratory distress syndrome | 0 | 1 (<0.1) | 0 | 0 | 0 | 1 (<0.1) |
| Arrhythmia | 1 (<0.1) | 1 (<0.1) | 0 | 1 (<0.1) | 1 (<0.1) | 2 (<0.1) |
| Myocarditis | 0 | 0 | 1 (<0.1) | 1 (<0.1) | 1 (<0.1) | 1 (<0.1) |
| Pericarditis | 0 | 0 | 0 | 0 | 0 | 0 |
| Bell's palsy | 1 (<0.1) | 0 | 1 (<0.1) | 0 | 2 (<0.1) | 0 |
| Facial Paralysis | 0 | 0 | 1 (<0.1) | 1 (<0.1) | 1 (<0.1) | 1 (<0.1) |
| Deep vein thrombosis | 0 | 0 | 1 (<0.1) | 0 | 1 (<0.1) | 0 |
| Embolism | 0 | 0 | 0 | 0 | 0 | 0 |
| Cerebral venous sinus thrombosis | 0 | 0 | 0 | 0 | 0 | 0 |
| Thrombocytopenia | 0 | 0 | 0 | 0 | 0 | 0 |
| Disseminated intravascular coagulation | 0 | 0 | 0 | 0 | 0 | 0 |
| Guillain-Barré syndrome | 0 | 0 | 0 | 0 | 0 | 0 |
